# Potential health and cost impacts of a point-of-care test for neonatal sepsis and possible serious bacterial infections in infants: a modeling analysis in two settings

**DOI:** 10.1101/2024.12.03.24318382

**Authors:** Joshua M Chevalier, Megan A Hansen, Kyra H Grantz, Birgitta Gleeson, Benjamin Blumel, Veronicah Chuchu, Shaukat Khan, Ntombi Sigwebela, Gwendoline Chimhini, Felicity Fitzgerald, Cecilia Ferreyra, Brooke E Nichols

## Abstract

**Background:** Sepsis contributes to nearly 50% of neonatal deaths in resource-limited settings. Accurate, timely diagnosis can improve outcomes, reduce inappropriate antibiotic use, and save healthcare costs. We aimed to determine the minimum technical requirements and cost of a point-of-care test (POCT) for neonatal sepsis to be clinically effective in hospitals and community settings in low-resource settings.

**Methods:** We modeled the diagnosis and treatment of hospitalized neonates and infants at primary care facilities with suspected sepsis. Health outcomes (mortality, hospital stays, and healthcare-associated infections (HAIs)) were compared under empiric treatment with varied blood culture scenarios to a POCT. Test performance, bacterial infection prevalence, and discharge criteria were varied. A threshold economic analysis identified maximum allowable costs for a cost-neutral POCT.

**Results:** A POCT could reduce neonatal deaths significantly at hospitals (up to 19%) and community levels (up to 70%), compared to baseline, by enabling faster therapy initiation and reducing unnecessary hospitalizations and HAIs. Healthcare costs could drop by 17%-43% in hospitals and 48%-81% in primary care settings. A POCT priced at $21 for hospitals and $3 for community use could remain cost neutral.

**Conclusions:** A POCT for neonatal sepsis, even with moderate accuracy, could improve outcomes by accelerating diagnosis, supporting antibiotic stewardship, and lowering costs. High sensitivity is essential to minimize deaths from missed diagnoses and delayed antibiotic therapy. Our results suggest 85% sensitivity and 80% specificity for hospitalized neonates and 90% sensitivity, and 70% specificity in primary care settings are the minimum necessary technical requirements for the POCT.

## Introduction

Sepsis is a significant cause of neonatal mortality worldwide, accounting for 7.3% of neonatal deaths, disproportionately affecting low-and middle-income countries (LMICs) [1,2].

Diagnosing neonatal sepsis is challenging due to nonspecific symptoms (i.e., fever, irritability) and limited diagnostic resources, particularly in LMICs where blood culture services are often unavailable or unreliable [3]. Without timely and adequate diagnosis, neonates or infants suspected of sepsis are empirically treated with broad-spectrum antibiotics, leading to antibiotic overuse, increased resistance, and unnecessary hospitalization of those without bacterial infections. Antibiotic overuse in this age group can lead to morbidity and mortality, especially in low birthweight or preterm newborns, while also contributing to the growing concern of antimicrobial resistance (AMR) [4,5]. Continued hospitalization of otherwise healthy neonates puts them at risk of healthcare-associated infections (HAIs), a major cause of sepsis [6].

Likewise, in community-level care, no diagnostic tests are available to diagnose infants with possible serious bacterial infections (pSBI). Infants are typically diagnosed clinically, treated with a single dose of antibiotics, and referred to hospitals [7,8]. If referral is not feasible due to distance, cost, and/or time required, antibiotic injections may be provided at the PHC level [9].

New diagnostic tools to quickly identify neonatal sepsis and pSBI could improve care, guide referrals, and reduce misdiagnosis [7,10]. Current biomarker candidates for a point-of-care test (POCT) include C-reactive protein (CRP), procalcitonin (PCT), or Serum Amyloid A (SAA) [11–13]. Clinical algorithms or prediction scores have also been considered [14]. A POCT for neonatal sepsis must attain certain technical specifications (sensitivity/specificity) and a realistic price to be clinically useful and feasibly implemented [7,15]. To understand these minimum diagnostic criteria and their impact on health-outcomes, we modeled clinical cascades of neonates born in-hospital and infants presenting at PHC facilities suspected of sepsis with current standard-of-care, or a POCT with same-day results. We performed a threshold economic analysis to determine the prices at which POCT implementation would be cost neutral to the health system.

These results supported the development of a Target Product Profile (TPP) for POCTs for neonatal sepsis and pSBI in infants. The TPP identifies two priority use-cases: 1) in hospitals, when newborns are being evaluated for sepsis, and 2) in community (non-hospital) settings, where infants are evaluated for pSBI. The TPP provides guidance for regulatory authorities, manufacturers, ministries of health, and other health programs and stakeholders for test development [10].

## Methods

To evaluate the potential impact of a true POCT for neonatal sepsis, we deterministically modeled the diagnosis and clinical care cascade of two cohorts: 1) hospital-born neonates 0-28 days old and 2) infants 0-59 days old presenting for care at the PHC level. Both models are static and parameterized using point estimates from studies in India, Uganda, or other representative countries with available data (**Table 1**) (**Supplement, Text S1-S3)**. Models were built in RStudio 4.3.1, and analyses were conducted using RStudio and Microsoft Excel 16.81.

**Table 1.** Model parameters for hospital and community settings. *\*CFR: Case Fatality Rate*.

| Parameter | Value (%) |
| --- | --- |
| **Hospital Model** |  |
| Prevalence of culture-positive sepsis among suspected cases [21,28] |  |
| - *High* | 23.6 |
| - *Moderate* | 11.8 |
| - *Low* | 5.9 |
| Proportion of infections which are antimicrobial resistant [3] | 50.0 |
| Proportion of suspected cases receiving blood culture [21] | 59.5 |
| Blood culture sensitivity/specificity [29] | 66.7/100 |
| Probability of antibiotic-related morbidities among antibiotic-treated children [29] | 27.7 |
| Probability of acquiring hospital-associated infection during hospitalization among children without prior infection [30] | 5, 10, 15, 20 |
| CFR*, no infection [3] | 8.8 |
| CFR*, antibiotic susceptible infection [3] | 29.0 |
| CFR*, antimicrobial resistant infection [3] | 54.5 |
| CFR*, delayed antibiotic initiation [17] | 68.2 |
| CFR*, antibiotic-associated mortality [31] | 0.9 |
| **Community Model** |  |
| Possible serious bacterial infection [8,9] |  |
| - *Proportion fast breathing* | 38.7 |
| - *Proportion clinical severe infection* | 53.9 |
| - *Proportion critical illness* | 7.3 |
| Bacterial prevalence [3,21,28] |  |
| - *Weighted average (0.5x, 1.0x, 2.0x)* | 5.2, 10.4, 20.8 |
| - *Fast breathing (0.5x)* | 3.0, 5.9, 11.8 |
| - *Clinical severe infection (1x)* | 5.9, 11.8, 23.6 |
| - *Critical illness (2x)* | 11.8, 23.6, 47.2 |
| CFR* (community-treated) [8] |  |
| - *Fast breathing* | 0.1 |
| - *Clinical severe infection* | 1.9 |
| - *Critical illness* | 14.6 |
| CFR* (hospital-treated) [8,21,32] |  |
| - *Non-bacterial illness* | 2.6 |
| - *True bacterial infection* | 11.5 |
| - *Healthcare-associated infection* | 12.0, 29.0 |
| Referral acceptance [8,9] | 17, 37, 69, 90 |
| **Cost Assumptions** |  |
| Daily cost of treatment and care in tertiary hospital (India) [24,25] |  |
| - *Non-bacterial illness* | \$96 |
| - *True bacterial infection* | \$118 |
| Daily cost of treatment and care in tertiary hospital (Uganda) [25,26] |  |
| - *Non-bacterial illness* | \$31 |
| - *True bacterial infection* | \$39 |
| Cost of oral amoxicillin (India) [24,25,27] |  |
| - *Community-treated* | \$1.29 |
| - *Hospital-treated* | \$1.88 |
| Cost of oral amoxicillin (Uganda) [25,26] |  |
| - *Community-treated* | \$0.43 |
| - *Hospital-treated* | \$0.62 |
| Cost of outpatient visit to hospital after referral (India) [25,27] | \$18.16 |
| Cost of outpatient visit to hospital after referral (Uganda) [25,26] | \$5.99 |

### Cohort 1: Hospitalized neonates

The modeled 1,000 hospital-born neonates (0-28 days) suspected of sepsis due to risk factors (early gestational age, low birthweight) or clinical signs indicative of sepsis (fever, difficulty breathing, inconsistent heart rate). Varied parameters included true bacterial prevalence and antibiotic resistance rates, which increased case fatality rates (CFRs) (**Table 1**). **Figure 1A** displays the clinical care cascade of the modeled population. Baseline care involved empiric antibiotic therapy and partial blood culture access, with delayed results (48 hours) reducing clinical utility.

**Figure 1.**
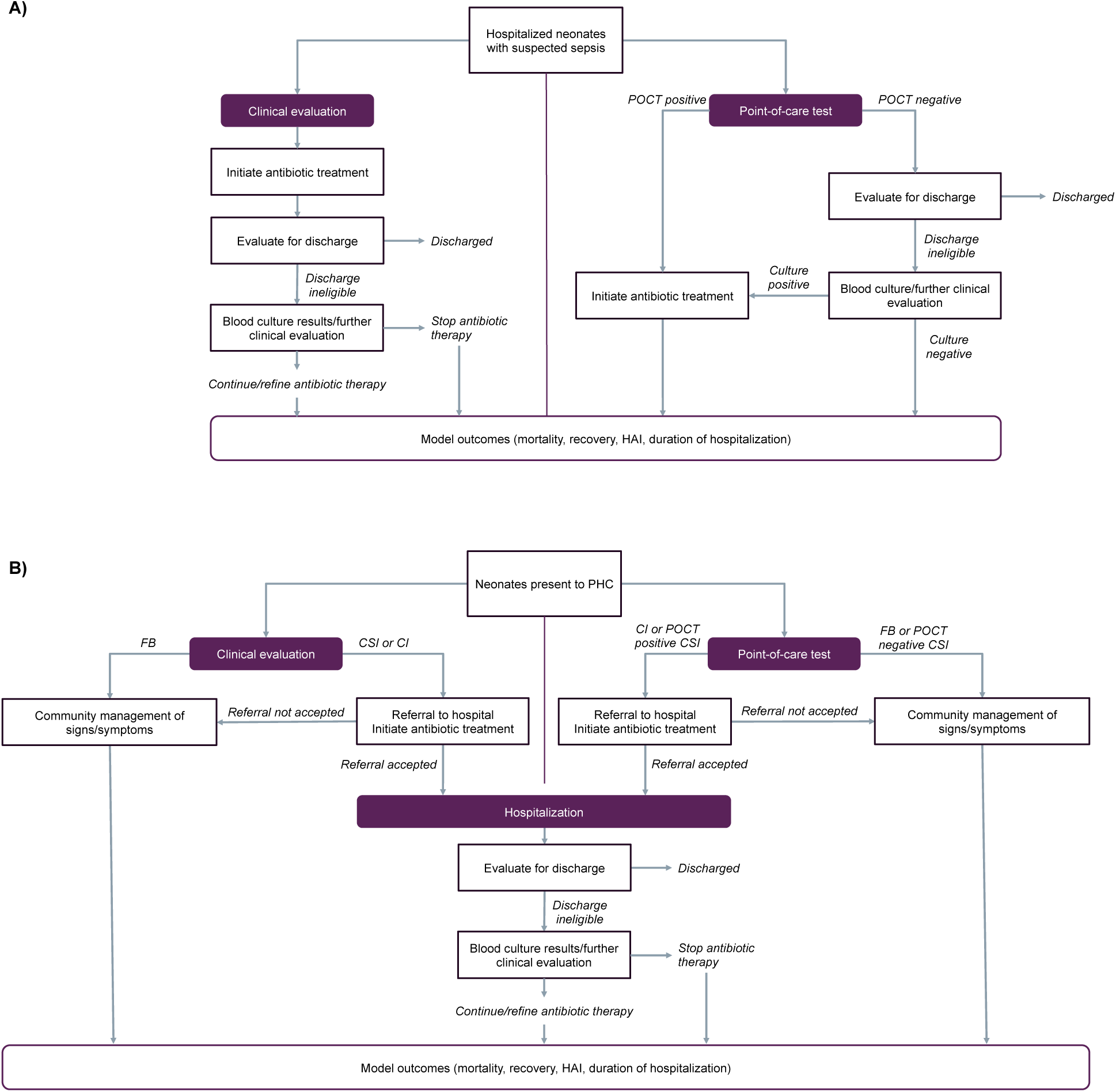
Conceptual diagram for clinical cascade for a point-of-care test for neonatal sepsis at the primary health care level and hospital. *\*POCT: point-of-care test, PHC: primary health care, FB: fast-breathing, CSI: clinical severe infection, CI: critical illness, HAI: hospital-associated infection*

Under POCT intervention, all neonates with suspected sepsis were tested using the POCT, and only those with a positive result immediately initiated antibiotics, allowing for potential early discharge for neonates testing negative. We considered varying proportions of neonates without true bacterial sepsis eligible for discharge after clinical evaluation and a negative result (blood culture or POCT) to represent well-presenting neonates. Discharge eligibility ranged (23%, 35%, 58%) based on the number of neonates discharged within two, three, or five days from a neonatal intensive care unit (NICU) [16]. We assumed neonates with a false-negative result remained hospitalized based on their clinical presentation and received additional diagnostic testing. This created a delay to antibiotic therapy (>0.5 hours), resulting in two additional days spent in the hospital and a 47% increase in the CFR [17,18]. Neonates without a true bacterial infection who received antibiotic therapy from empiric treatment or false-positive results and remained hospitalized were at risk of antibiotic-related morbidities and HAIs. Neonates with a true bacterial infection (true positive) had an average 16-day hospital stay, while neonates without infection (false positive) stayed an average of seven days [16]. Neonates with an HAI spent 11 additional days in the hospital, for a total 18-day stay [19].

### Cohort 2: Infants in primary health care facilities

We modeled 5,000 infants (0-59 days of age) presenting to PHC facilities with pSBI, categorized by illness severity (**Table 1)** [8,20]. Each pSBI category had an assumed underlying probability of true bacterial infection, which was varied in this analysis. Bacterial prevalence was parameterized lower in fast breathing infants and higher in critically ill infants, compared to infants with clinical severe infection, reflecting correlation between pSBI severity and true bacterial infection [21].

Under the standard-of-care (WHO guidelines) fast breathing infants 7-59 days of age were treated on-site with oral amoxicillin, while infants with clinical severe infection and critical illness were referred to inpatient care with a pre-referral dose of antibiotics [22] (**Figure 1B**). When referral was not feasible for infants with clinical severe infection or critical illness, we assumed they were treated on-site for up to seven days with WHO-recommended antibiotic regimens. The probability of accepting referral to hospitalization (17%-90%) was varied to reflect differently resourced contexts [8]. CFRs for each pSBI category were derived from the literature (**Table 1**).

Using the POCT, all infants presenting with critical illness received a referral and a single antibiotic dose regardless of their POCT result, while all fast-breathing infants were treated on-site without referral based on their POCT result. Infants presenting with a clinical severe infection were only referred for higher-level care, with one dose of antibiotics, if their POCT was positive; otherwise, those with a negative POCT remained monitored on-site. False positives with clinical severe infection received referral and treatment, while false negatives had an assumed four-hour delay to antibiotic initiation and increased CFR [17,23].

Upon referral acceptance, infants entered the hospital model (**Figure 1A**) without additional testing using the POCT. Hospitalized infants without a true underlying bacterial infection were at risk of HAIs, experiencing a higher CFR compared to community-borne infections. These CFRs differed from those in the hospital model because different clinical populations are considered in each model (**Table 1**). Discharge eligibility among community-referred infants without bacterial infection was held constant at 58%.

### Outcomes and Sensitivity Analysis

Diagnostic impact was quantified as the percent reduction in deaths and hospital days, and the number of HAIs and antibiotic-associated morbidities prevented through POCT use. Diagnostic impact was assessed across a range of POCT sensitivities (80%-100%) and specificities (70%-100%). Sensitivity analyses were performed to understand the performance of the POCT across several scenarios: varying discharge eligibility, referral acceptance, true bacterial prevalence among pSBI cases (6%, 12%, 24%), and the risk of contracting an HAI (5%-20%). In the community-level analysis, the CFR of HAIs was varied (12%, 29%).

### Economic Analysis

A threshold economic analysis evaluated the total costs of care at baseline and with the POCT to determine at which price the POCT would be cost neutral to the healthcare system including direct and indirect costs (i.e., training, supply chain logistics, quality control), a key characteristic of the TPP. Total inpatient care costs in India and Uganda, representing variation, were gathered from the provider perspective and inflated to 2023 USD (Table 1) [24–26].

Healthy infants did not contribute additional costs after discharge. The maximum allowable cost for a POCT in a hospital setting was calculated by dividing hospitalization costs saved using the POCT by the total POCTs administered to achieve cost savings.

The maximum allowable cost of a POCT at PHCs was also determined using cost savings of community management and administered antibiotics (Table 1) [24–27]. Inpatient care costs at a hospital after referral followed those described for the hospital model. Healthcare costs in Uganda, where community care and referral costs were reduced by 33%, were used as a sensitivity analysis to estimate the maximum cost at which a POCT remained cost-neutral [26]. This analysis was also performed excluding infants with critical illness, as their POCT results did not alter clinical practice in the model.

## Results

### Health outcomes of hospitalized neonates following POCT implementation

In the hospital cohort, all neonates suspected of sepsis received empiric antibiotic therapy and 60% received blood culture. Blood culture results permitted early discharge of 5%-13% of neonates without true, depending on underlying eligibility (23%-58%). The final mortality rate among neonates who had or developed infection was 41.7%.

The percentage of neonatal deaths prevented with a POCT was greater with lower underlying bacterial prevalence (6%) and larger proportions of neonates eligible for discharge (58%). In these contexts, test specificity was particularly important to rule out infection among well-presenting neonates. Increasing test specificity reduced HAIs due to earlier discharge of neonates without infection and greater reductions of antibiotic-associated morbidities from fewer false-positive results (**Supplement, Figure S1-S2**). The impact of the POCT increased when HAI risk was higher (20% vs. 5%), as HAIs constituted 28%-66% of total infection related deaths with high risk versus 9%-33% when risk was low (**Figure 2**).

**Figure 2.**
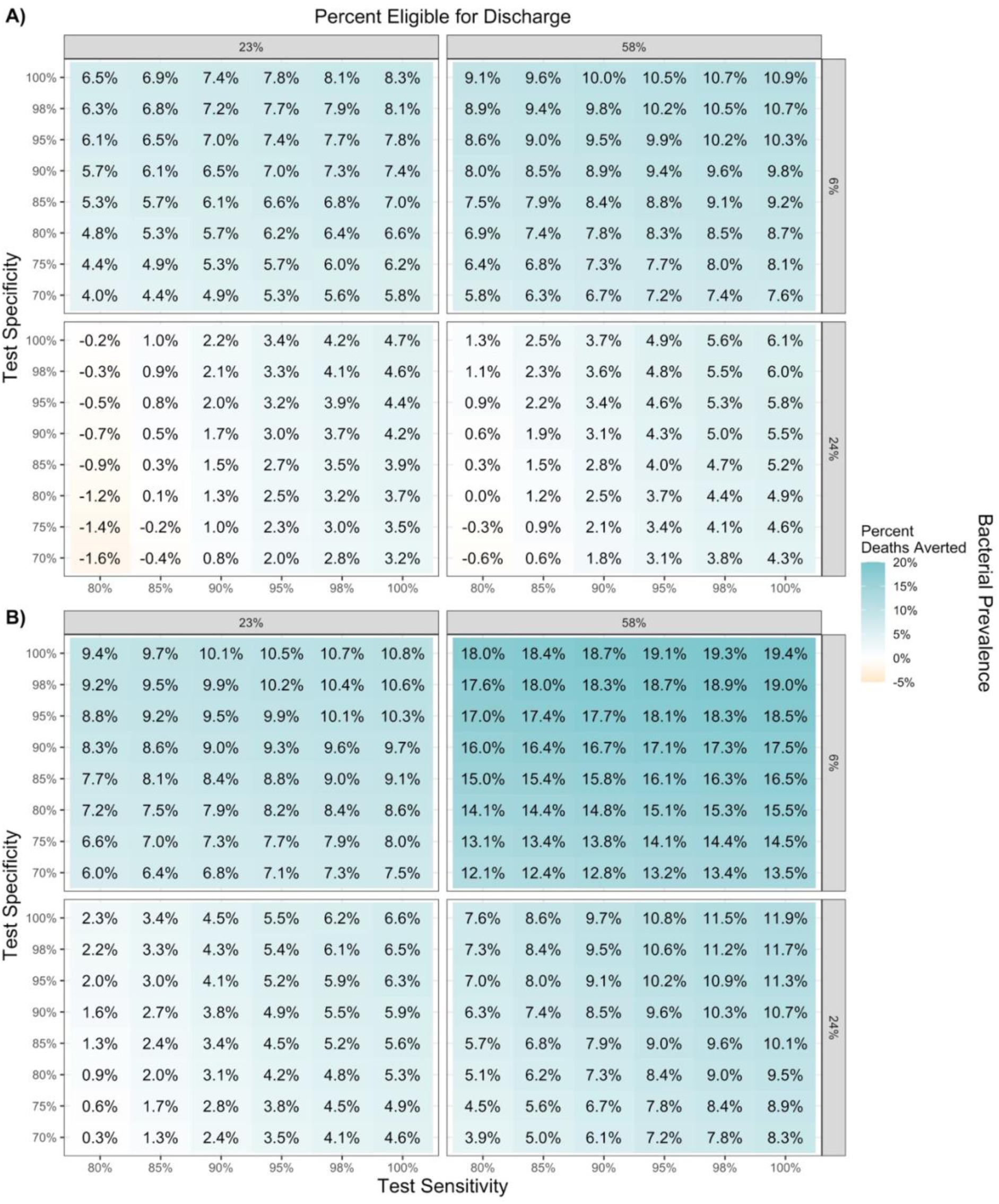
Percent reduction in neonatal deaths in a hospital setting, across modeled bacterial prevalence and proportion of neonates eligible for discharge, by test specificity and sensitivity, when **A)** the risk of hospital-associated infections is low (5%) or **B)** high (20%).

Yet, in some contexts test sensitivity was the key driver of diagnostic impact. In settings with low HAI risk (5%) and discharge eligibility (23%), but high bacterial prevalence (24%), excess deaths occurred with the POCT compared to baseline (0.2%-1.6% relative increase) when sensitivity was below 85% due to increasing false-negative results and subsequent delays in antibiotic therapy. With 85% sensitivity, a test specificity greater than 80% was required to avoid increased mortality across all contexts (**Figure 2A**).

With high HAI risk (20%), a POCT for neonatal sepsis in hospitals led to a net reduction in neonatal deaths (0.3%-19.4% of deaths averted compared to baseline) at all test sensitivities (80%-100%) and specificities (70%-100%) under the varying modeled proportions of bacterial prevalence and discharge eligibility (**Figure 2B**). The percentage of neonatal deaths prevented increased as POCT sensitivity increased, limiting false negatives and reducing delays to antibiotic initiation for infected neonates.

Earlier discharge leading to reduced HAI opportunity, produced a net reduction in hospital days. Total hospitalization days dropped by 8.2%-50.9% with POCT use **(Supplement, Figure S3**).

The percent reduction in hospital days followed similar trends to neonatal deaths, with respect to test specifications, discharge eligibility, and bacterial prevalence, but was not significantly impacted by HAI risk.

### Health outcomes of community-presenting infants following POCT implementation

In the modeled PHC cohort of 5,000 infants with pSBI, the proportion of infants correctly referred to upper-level care at baseline (i.e., infants with true bacterial infection) varied from 17%-33% as underlying bacterial prevalence increased (6%-24%). The proportion of correct referrals increased from 17% at baseline to 38% upon POCT implementation with the lowest modeled test specifications (80% sensitivity, 70% specificity) when bacterial prevalence was low and from 33% to 58% when bacterial prevalence was high (**Supplement, Figure S4**). The relative impact of the POCT on appropriate referrals largely relied on high test specificity prompting more correct referrals.

At baseline, 13-50 infant deaths occurred in PHC clinics and hospitals per 1,000 pSBI episodes across all modeled scenarios (**Supplement, Figure S5**). Mortality increased as underlying bacterial prevalence and referral acceptance increased. Greater referral acceptance led to increased hospital admission of infants without bacterial infection, posing HAI risk. HAI deaths composed between 0.6%-44.7% of total deaths at baseline, depending on other varied parameters (**Supplement, Figure S6**).

A POCT informing hospital referrals reduced total infant deaths when test sensitivity and specificity were sufficiently high (**Figure 3**). The range of scenarios in which deaths increased was greater when HAI risk and mortality were low (5%, 12% respectively) (**Figure 3A**).

**Figure 3.**
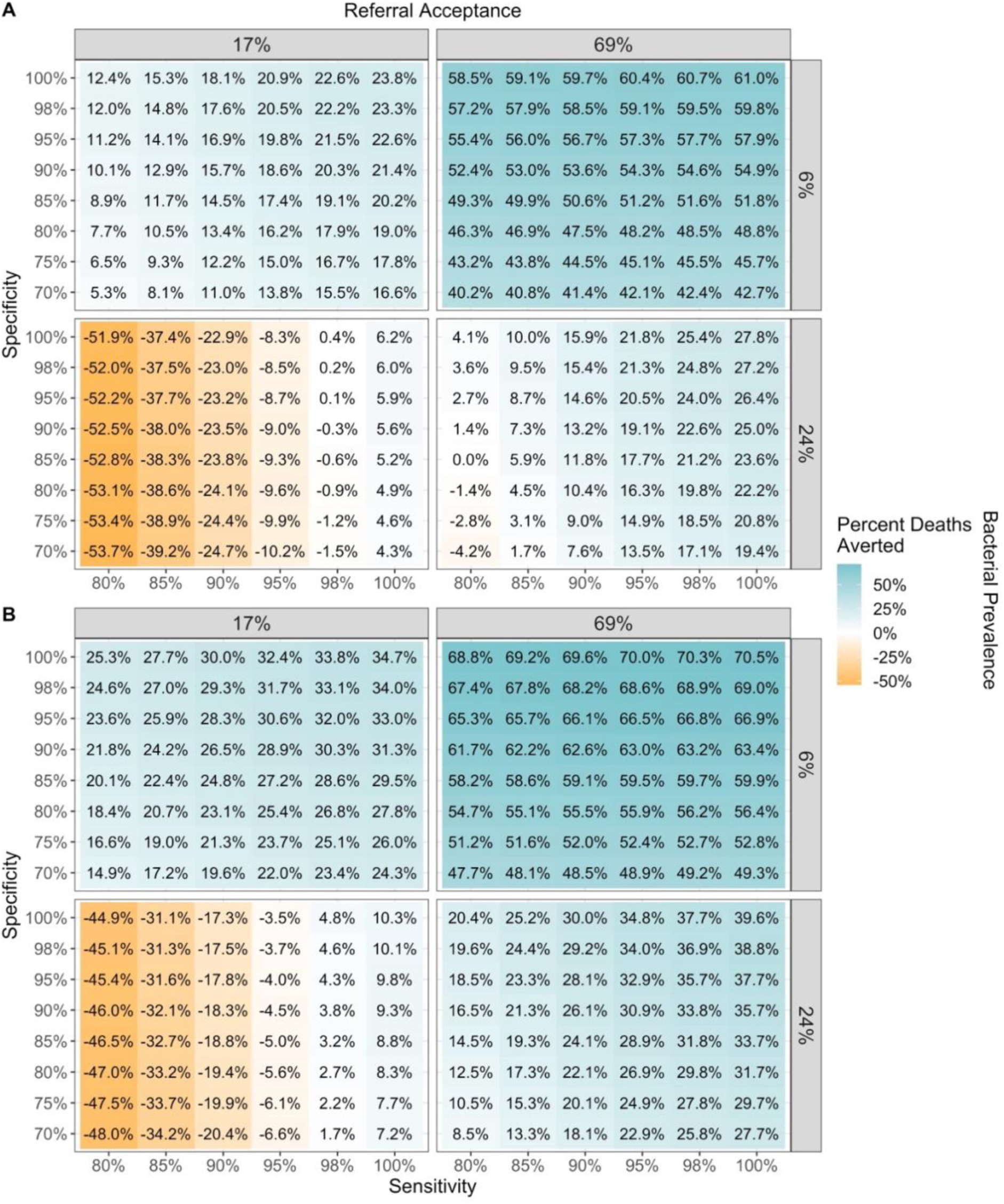
Percent of deaths averted compared to baseline when using the point-of-care test at the community level, by test sensitivity, specificity, bacterial prevalence, and referral acceptance, when A) HAI risk and mortality are low (5%, 12%), and B) HAI risk and mortality are high (20%, 29%)

At low bacterial prevalence and high referral acceptance, and high HAI risk (20%) and mortality (29%), a POCT with high sensitivity and specificity could reduce infant deaths by up to 70% (**Figure 3B**). However, with low sensitivity and specificity, more deaths were observed (3.5%-48%) with lower referral acceptance and higher levels of bacterial prevalence. This was attributable to delayed antibiotic initiation associated with a false-negative POCT.

To limit deaths caused by false-negative results, a high sensitivity of at least 90% could be combined with a lower specificity (70%), reducing total infant deaths across most scenarios. However, under the worst-case scenarios, test sensitivity (≥98%) and specificity (≥95%) would need to be exceedingly high to avoid excess infant mortality.

## Sensitivity Analysis

Univariate sensitivity analyses of parameters in both the hospital and community models, and their relationship with the number of infant deaths averted, stratified by HAI risk, are shown in **Figure 4**. Sensitivity and specificity remained at 85% and 80% in the hospital model, and 90% and 70% in the community model, representing the identified necessary minimum specifications to reduce mortality across most scenarios.

**Figure 4.**
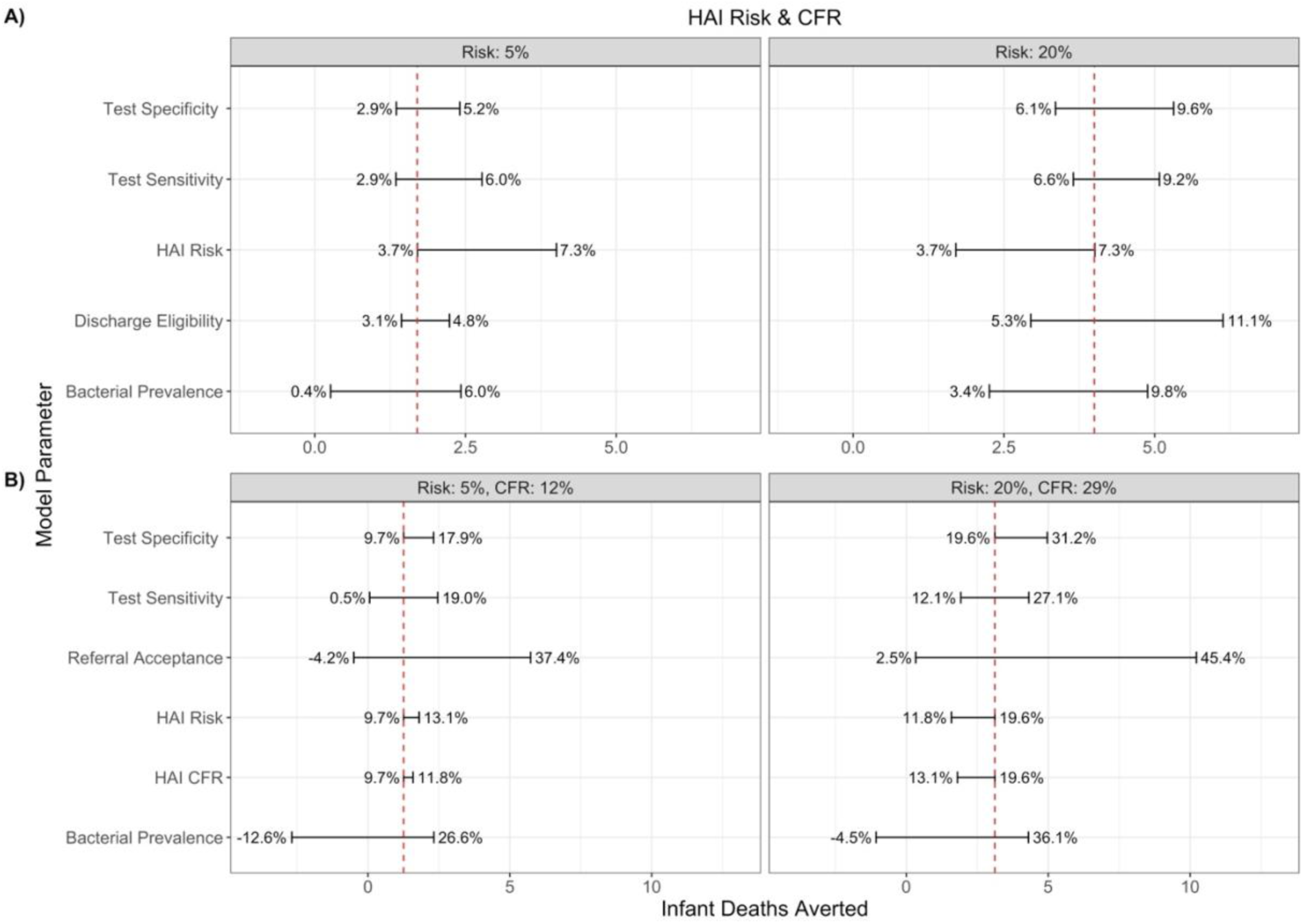
Univariate sensitivity analysis of A) hospital model parameters or B) community model parameters and the correlated number of neonatal deaths averted, stratified by A) hospital-associated infection risk or B) hospital-associated infection risk and case fatality rate. The absolute number of deaths averted is shown on the X-axis, while the corresponding percentage is shown on the error bar label. The vertical red, dashed line represents the number of deaths averted when all other variables are held at their constant A) specificity: 80%, sensitivity: 85%, discharge eligibility: 35%, bacterial prevalence: 12% or B) specificity: 70%, sensitivity: 90%, referral acceptance: 37%, bacterial prevalence: 12%. *\*HAI: hospital-associated infection, CFR: case fatality rate*

At the hospital level, increasing test specificity, promoting accurate negative diagnoses, drove reductions in neonatal deaths by earlier discharge and reducing HAIs. HAI risk substantially influenced the potential reductions in mortality; when HAI risk was low, there was less potential for the POCT to prevent deaths by eliminating unnecessary hospitalization.

At the community level, the probability of referral acceptance overwhelmingly influenced the impact of a POCT, dictating the possibility for accurate negative POCT diagnoses to avert unnecessary hospitalization. Overall bacterial prevalence also shaped POCT impact, with greater potential impact when prevalence was lower (i.e., when there were more healthy infants benefitting from accurate non-referral).

### Economic Analysis

POCT implementation in hospitalized neonates reduced hospitalization days (**Supplement, Figure S7**), cutting total care and treatment costs by 17%-43% at various levels of bacterial prevalence, proportion of neonates eligible for discharge, and test performance. Greater cost savings occurred at lower bacterial prevalence (6%) than higher prevalence (24%), highlighting the value of identifying discharge-eligible neonates without infection with a POCT (**Supplement, Figure S8-S9**). Cost savings did not perfectly mirror mortality trends **(Figure 2)** as higher mortality led to fewer hospitalization days and thus lower costs.

Cost savings were more considerable following POCT implementation at the community level (48%-81%) mainly by minimizing inappropriate referrals and hospitalization days. Fewer inappropriate referrals occurred at baseline when bacterial prevalence was low, granting more cost savings with POCTs. Across all levels of prevalence and discharge eligibility, cost savings increased with better test performance (higher sensitivity and/or specificity) (**Supplement, Figure S10-S11**). Excluding critically ill infants from the analysis raised the median allowable POCT cost, from $0.86-$1 in India and $0.28-$0.33 in Uganda, across varied scenarios (**Supplement, Figures S12-S13**).

To ensure the financial feasibility of implementing POCTs across all settings and ranges of uncertainty, our analysis indicates that a globally acceptable neonatal sepsis POCT in hospital settings should not exceed $21 USD. Likewise, to ensure cost neutrality in community settings, the lowest of costs saved per test administered ($3) should be selected as the optimal cost at PHC levels (**Supplement, Table S1-S2**). Results are reported following the Consolidated Health Economic Evaluation Reporting Standards (**Supplement, Table S3**).

## Discussion

Implementation of a sufficiently accurate POCT for sepsis detection in neonates or infants at hospital and community-level facilities can reduce overall morbidity and mortality and remain cost neutral when priced appropriately [10,14,33]. Although the exact impact of a POCT heavily depends on the specific use case implementation context, our modeling finds a test with at least 85% sensitivity and 80% specificity for hospitalized neonates and 90% sensitivity and 70% specificity for community-presenting infants is likely to reduce mortality.

If test sensitivity is lower than these values, deaths caused by missed diagnosis of true sepsis cases and ensuing antibiotic delays could increase overall mortality. However, there were instances where tests with low sensitivity and high specificity, due to large reductions in unnecessary hospitalization, antibiotic use, and HAI acquisition among uninfected patients, could reduce mortality and costs despite greater false-negative diagnoses. While the overall benefits of a POCT may outweigh the consequences of false-negative results in these scenarios, a test sensitivity well above 80% should remain the criterion, with a sensitivity of 90% serving as an ideal TPP standard. A POCT with low sensitivity would have limited utility in independently ruling out sepsis among neonates and infants.

Test specificity must also be sufficiently high to reduce unnecessary hospitalizations and HAIs. However, even a test with lower specificity will improve upon the standard-of-care, possibly tolerated in some contexts. In hospital settings, 80% specificity paired with 85% sensitivity generated a net reduction in deaths. Yet, in community settings, 70% specificity with 90% sensitivity reduced infant deaths in most scenarios, excluding contexts with low referral acceptance (17%) and high bacterial prevalence (24%). Under these circumstances, even 100% specificity is insufficient unless sensitivity reaches 98%, underscoring the need to evaluate contextual factors when implementing POCTs.

Given the complexity in diagnosing sepsis in neonates and infants, a POCT may be used as part of a diagnostic package, affecting overall diagnostic impact and modifying the requirements and trade-offs between POCT sensitivity and specificity. A true POCT may be hard to develop, but a near POCT could have less impact than modeled due to delayed results. The current biomarker candidates for POCTs, including CRP, PCT, and SAA have all shown sensitivities below the 80% threshold identified in this analysis [12,34,35]. The POCT could be part of a clinical algorithm incorporating signs, symptoms, and risk factors, lowering the required test performance for net benefit. We also did not account for how positive POCT results might improve antibiotic timing and outcomes beyond empiric therapy, suggesting additional uncaptured benefits.

A key limitation of this analysis is that model parameters, assumptions, and costs were all derived from tertiary hospital and PHC studies in India and Uganda and thus may not perfectly represent other resource-limited settings. Blood culture was assumed to be 100% specific in diagnosing true negatives, but culture contamination is common in these settings creating false-positive results, potentially altering health outcomes. Likewise, the possibility of false-negative results due to inadequate blood volume for optimal culture performance could also impact our estimates. The model contains assumptions that may differ across diverse environments where sepsis in neonates and infants is common, including bacterial prevalence, resistance profiles, length of hospitalization, and HAI rates. To account for this variability, several parameters were varied to improve model sensitivity and observe trends to inform the suggested values to be included in the TPP for sensitivity, specificity, and costs. While we expect relative differences between the POCT and baseline standard-of-care to be consistent, these results should be interpreted with discretion as parameters continue to evolve with potential stock-outs of appropriate antibiotic treatment and exponential AMR spread in resource-limited settings.

Additionally, simplifying assumptions regarding clinical management and outcomes were made, which may not capture all challenges of clinical pathways in sepsis diangosis. For example, infants may become lost to follow-up after PHC referral, incurring unknown health outcomes.

Patients were functionally dichotomized into those who were being appropriately treated with antibiotics and untreated; in reality, treatment is complex, and there are clinical benefits to continued antibiotic refinement that could not be incorporated here. Similarly, indirect long-term benefits to implementing POCTs that exist are not explicitly modeled here, including reductions in AMR spread and preservation of first-line antibiotics in low-resource settings. Finally, we assumed the POCT performed as intended with respect to sensitivity and specificity but ignored possible invalid results occurring from new technology. Clarifying diagnostic algorithms based on available and developing technologies, and the clinical management that accompany them are necessary for future accurate costing of these tools – especially as practical challenges (i.e., operational and supply chain procedures) are identified during prototype development and implementation [36].

This modeling analysis aims to guide understanding of necessary test specifications to be considered with other technical parameters (i.e., blood volume, sample preparation guidelines, and result timelines) as a call for the development of a POCT as an aid to diagnose sepsis in hospitalized neonates and community-presenting infants. An accurate POCT could improve time to diagnosis and generate net reductions in hospital days, HAIs, and deaths, while promoting the de-escalation of antimicrobials in resource-limited settings. Our results displayed a critical trade-off between HAI-related deaths prevented and those caused by a false-negative diagnosis, suggesting that a high sensitivity should be prioritized over specificity to avoid potential delays in antibiotic initiation. Given the nature of the settings in which the POCT will be used, its cost should not exceed $21 in hospital settings and $3 in community settings.

Future research should address the current gaps in data on the incidence and clinical management of sepsis in both hospital and community settings [37]. Bridging this gap is essential to generate robust, context-specific estimates of the impact of a POCT and its downstream effects on AMR [38]. Key areas of investigation may include advancing blood culture capacity, expanding polymerase chain reaction diagnostics and their accessibility, and validating the use of biomarkers. Nonetheless, urgent investments in diagnostic development are needed to accelerate access to diagnostic solutions for the vulnerable populations most impacted by neonatal sepsis.

## Author Contributions

Conceptualization: BG, CF, BEN. Data curation: JMC, MAH. Methodology: JMC, MAH, KHG, SK, BEN. Formal analysis: JMC, MAH, KHG. Investigation: JMC, MAH, BG, BB VC, KHG. Supervision: BG, CF, BB, SK, BEN, KHG. Writing - original draft: JMC, MAH. Writing - review and editing: JMC, MAH, KHG, BG, BB, VC, SK, NS, GC, FF, CF, BEN.

## Data Availability

We acknowledge the importance of transparent and responsible data sharing to foster scientific collaboration and advance research. Code for the model is available using the following link: https://doi.org/10.6084/m9.figshare.c.7776176.v1.All data used for this study are presented in the manuscript and gathered from publicly available literature and therefore do not necessitate any information de-identification. For additional information, interested parties can contact JMC or MAH.

https://doi.org/10.6084/m9.figshare.c.7776176.v1.

## Acknowledgements

Not applicable.

## Patient Consent Statement

This research utilizes publicly available data from literature; therefore, patient consent and/or formal ethical approval was not required. However, ethical considerations were included throughout the research process as transparency was maintained in the reporting of our modeling methodology. This statement affirms our commitment to ethical conduct in research, even in the absence of formal approval.

