## Supplement for "Potential health and cost impacts of a point-of-care test for neonatal sepsis and possible serious bacterial infections in infants: a modeling analysis in two settings"

### Hospital Model Parameters

*Variables follow a normal distribution based on the mean and standard errors presented, unless otherwise specified.*

**Suspected Sepsis** [1]

Mean proportion: 32.58% (4408/13530)
Standard error: 0.71%

**Culture Positive Sepsis (of suspected sepsis cases)**

**High prevalence** [2] (Double moderate prevalence, assumed)
Mean proportion: 23.6%
Standard error: 0.36%

**Moderate prevalence** [2]
Mean proportion: 11.80% (964/8165)
Standard error: 0.36%

**Low prevalence** [3]
Mean proportion: 5.93% (27/455)
Standard error: 1.11%

**Proportion receiving blood culture** (of suspected sepsis cases) [2]

Mean proportion: 59.51% (4859/8165)
Standard error: 0.54%

**Blood culture sensitivity** [4]

*Standard blood culture methods as compared to automated blood culture (BACTEC). This does not consider an inability to draw blood from a neonate, which would reduce the sensitivity further.

Mean proportion: 66.67% (24/36)
Standard error: 7.86%

**Antimicrobial resistant bacterial sepsis**

**High** [5]
Mean proportion: 75.48% (N=10894, based on weighted percentages)
Standard error: 0.41%

**Moderate** [1]

Mean proportion: 49.96% (420/840)
Standard error: 1.61%

**Low** [2]
Mean proportion: 16.67% (17/102)
Standard error: 3.69%

__**Antibiotic related morbidities and mortality**__ [6]

*For neonates with a moderate antibiotic use rate (9–32% of hospital days on ABs; 2nd and 3rd quartiles in study)

**Morbidities** (including persistent periventricular echogenicity or echolucency on neuroimaging, stage 3 or higher ROP, and CLD.)

Mean proportion: 27.71% (1226/4425)
Standard error: 0.67%

**Mortality** (of those who received ABs without underlying bacterial infection, part of the composite study outcome. Mortality increased as AB-use quartile increased, indicating a possible association. The association with mortality has remained through multivariable analysis in similar studies.[7])

Mean proportion: 0.88% (39/4425)
Standard error: 0.14%

__**Length of stay in hospital**__ [8]

| Days | Non-Sepsis (proportion) | Sepsis (proportion) |
|---|---|---|
| <24 hours | 1.09% | 0% |
| 1 | 12.52% | 1.74% |
| 2 | 12.52% | 1.74% |
| 3–5 days | 37.55% | 5.23% |
| 6–10 days | 23.11% | 30.75% |
| 11–30 days | 12.53% | 56.42% |
| 31–60 days | 0.68% | 4.11% |

**Increased length of stay for POCT false negatives**
*Based on a study that saw a difference in the mean length of stay for neonates with drug-related problems. Here we consider the drug-related problem to be delay in antibiotic initiation [9].

Mean difference: 2.1 (10.1–8 days)
Standard error: 0.8647

**Proportion of well-presenting neonates**

*Based on the length of stay study referenced above, we calculated the proportion of neonates (without sepsis) who survived and were discharged within 2–5 days. We assume these infants are most likely to be well enough for early discharge after a negative sepsis diagnosis and represent those eligible for release.[8]

| Days | Surviving neonates discharged (%) |
|---|---|
| 2 | 23.44 |
| 3 | 34.99 |
| 5 | 58.08 |

**Risk of hospital acquired infection (HAI)** [10]

*Underlying risk of HAI on hospital ***day one***

Rate: 5, 10, 15, 20% [11]

Mean LOS: 7 (neonates without pre-existing bacterial infection)

Per day risk:

Probability of HAI <- HAI rate * (1- exp(-log(2)/mean LOS))

| HAI Rate | Per Day Probability of HAI |
|---|---|
| 5% | 0.47% |
| 10% | 0.94% |
| 15% | 1.41% |
| 20% | 1.89% |

**Increased length of stay for HAIs** [12]
11.2 days (95%CI: 9.1–13.2)
Standard error: 1.05

**Case fatality rates**

*CFR susceptible bacterial (normal)* [2]
Mean: 29.00%
SE: 1.5%

*CFR AMR bacterial (normal)* [1]
Mean: 54.51%
SE: 1.6%

*CFR HAI bacterial*
**Average of CFR susceptible and AMR for a given simulation*

*CFR bacterial antibiotic delay (normal)* [13]
**OR of 1.47 on mortality with delay to AB initiation per 30-minutes*

*CFR antibiotic-related (normal)* [6]
Mean: 0.88%
SE: 0.14%

**Community Model Parameters**

*Possible Serious Bacterial Infection (pSBI)*

pSBI lower bound: 7.57% [14]
pSBI upper bound: 12.94% [2]
**Uniform distribution*

*Level of severity of presenting case* [14]

Clinical severe infection (CSI): 53.94% SE: 0.58%
Critical illness (CI): 7.34% SE: 0.03%
Fast breathing only (FB): 38.72% SE: 0.56%

Bacterial prevalence by severity

CSI prevalence: 11.80%, SE: 1.02% [2]
CI prevalence: CSI prevalence * 2 (assumed)
FB prevalence: CSI prevalence *0.5 (assumed)

CFR by level of severity [14]

Clinical severe infection (CSI): 14.6% SE: 1.71%
Critical illness (CI): 1.90% SE: 0.23%
Fast breathing only (FB): 0.10% SE: 0.05%

CFR HAIs: (12%, 29%) [2,11]

CFR delay odds ratio per 30-minute delay: 1.47 [13]

- Assuming a 4-hour antibiotic delay

Referral acceptance [14]

Lower bound: 37.2% (India)
Upper bound: 69.2% (Ethiopia)

Four-day follow-up community treatment: 78.00%, SE: 3.38%
Seven-day follow-up community treatment: 39.19%, SE: 3.99%

### Model Assumptions

- All suspected sepsis cases will receive antimicrobials at baseline, and all suspected sepsis cases will receive a point-of-care test under the intervention

- Hospitalization is based on clinical signs of illness and not diagnostic results, although diagnostic results may influence time to discharge.

- Suspected sepsis cases are ill-presenting (having signs of possible serious bacterial infection) and therefore follow respective hospitalization pathways. Neonates with no underlying bacterial infection spend less time in the hospital on average.

- Negative blood culture results will lead to discharge for a proportion of neonates without underlying bacterial infection on day 2 of hospitalization (23–58%). In the non-bacterial hospitalization pathway, 2016 neonates out of 3471 who survived (58.08%), were released within 5 days of hospitalization, indicating less severe illness. It is these neonates who are assumed to be eligible for discharge on day 2. Additionally, we consider the study proportions discharged on days 2 (23.44%) and 3 (34.99%) to represent different proportions of well neonates.

- Point-of-care false negatives with true underlying bacterial infection follow the same hospitalization pathway as test-true positives but spend an average of 2 additional days in the hospital due to delay in antibiotic initiation.

- Out of point-of-care true-test negatives, 58.08% (23.44%, 34.99%) will be eligible for early release, while 41.92% (65.01%, 76.56%) are assumed to be severe enough to warrant continued hospitalization and further diagnostic testing (blood-culture).

- All point-of-care test positives in addition to the 41.92% (65.01%, 76.56%) of test-negatives undergo additional blood culture.

- Point-of-care false positives with no underlying infection follow the non-bacterial hospitalization pathway. The proportion of which who receive blood culture, out of those who may be eligible for early release based on clinical severity (23.44–58.08%), are discharged upon negative result.

- Clinical management of point-of-care true-test positives is considered to have a possible improvement on the fatality rate (0–20%).

**Figure S1.** Percent reduction in hospital-associated infections among neonates in a hospital setting, across different proportions of discharge eligibility with increasing test specificity.

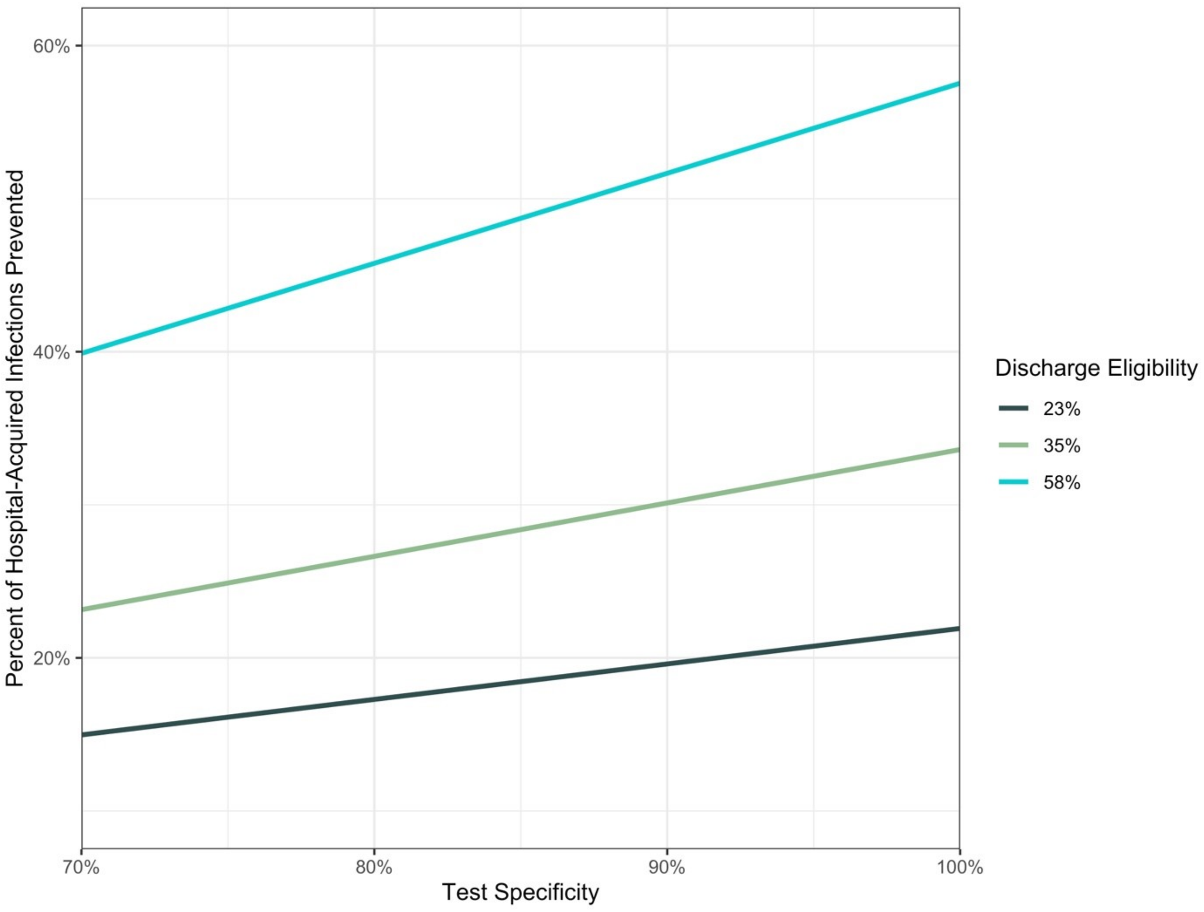

**Figure S2**. Antibiotic-related morbidities prevented per 100 suspected sepsis cases, in a hospital setting, with increasing test specificity.

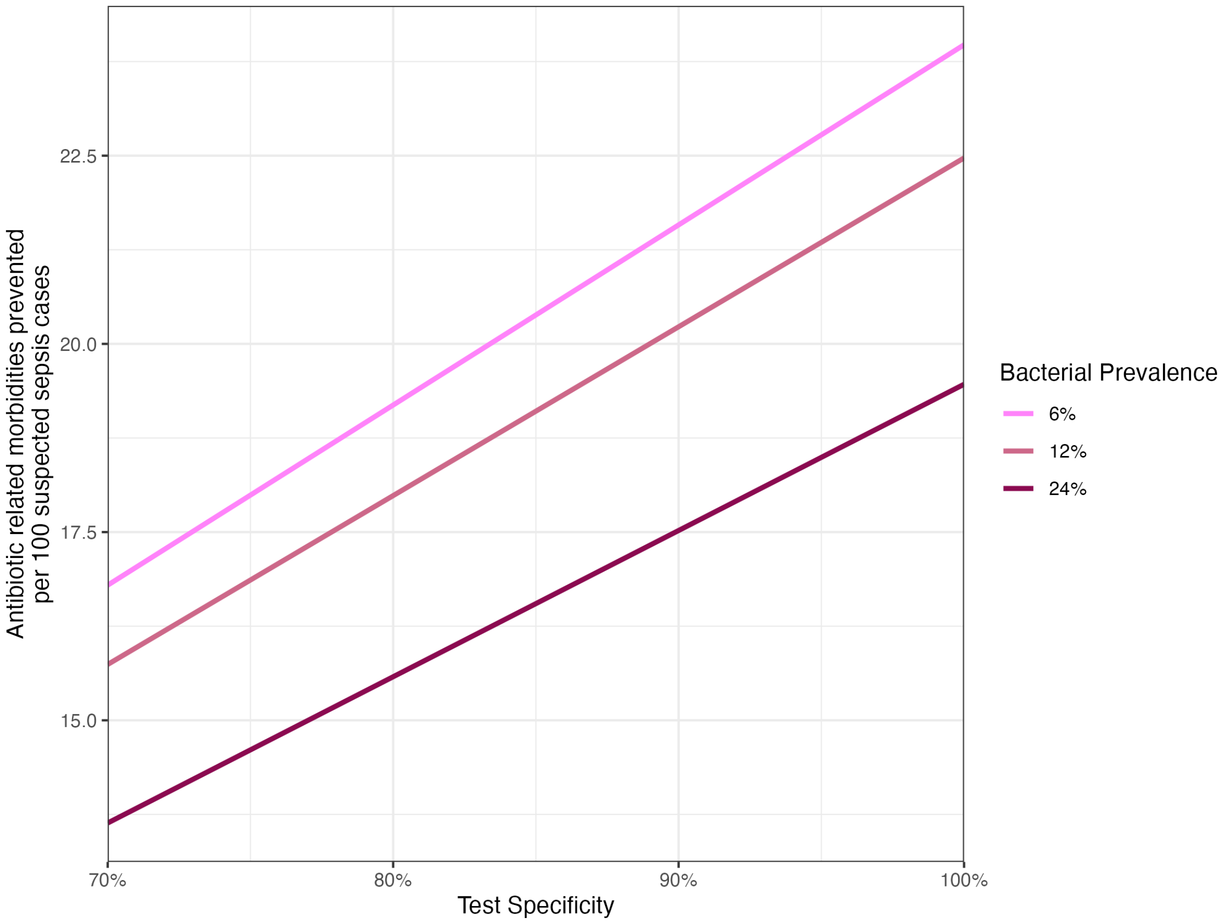

**Figure S3**. Percent reduction in hospital days across modeled bacterial prevalence and proportion of neonates eligible for discharge, by varying test specificity and sensitivity when A) the risk of hospital-associated infections is low (5%) or B) high (20%). **NICU: neonatal intensive care unit*

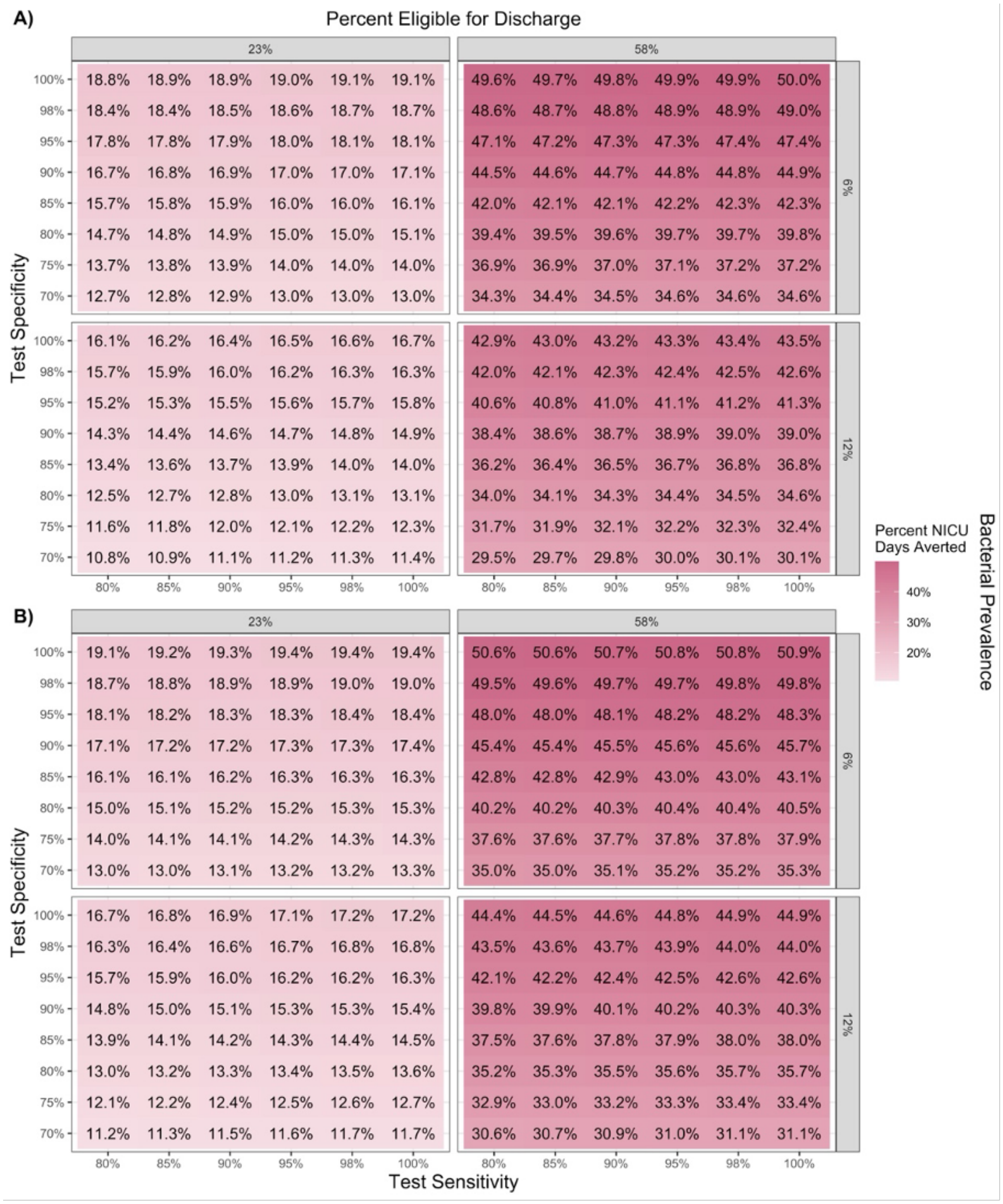

**Figure S4.** Percentage of correct referrals at baseline in the community setting, by test specificity and bacterial prevalence. The solid line represents the proportion of correct referrals with the point-of-care test and the dotted line represents the proportion of correct referrals at baseline, for the respective bacterial prevalence.

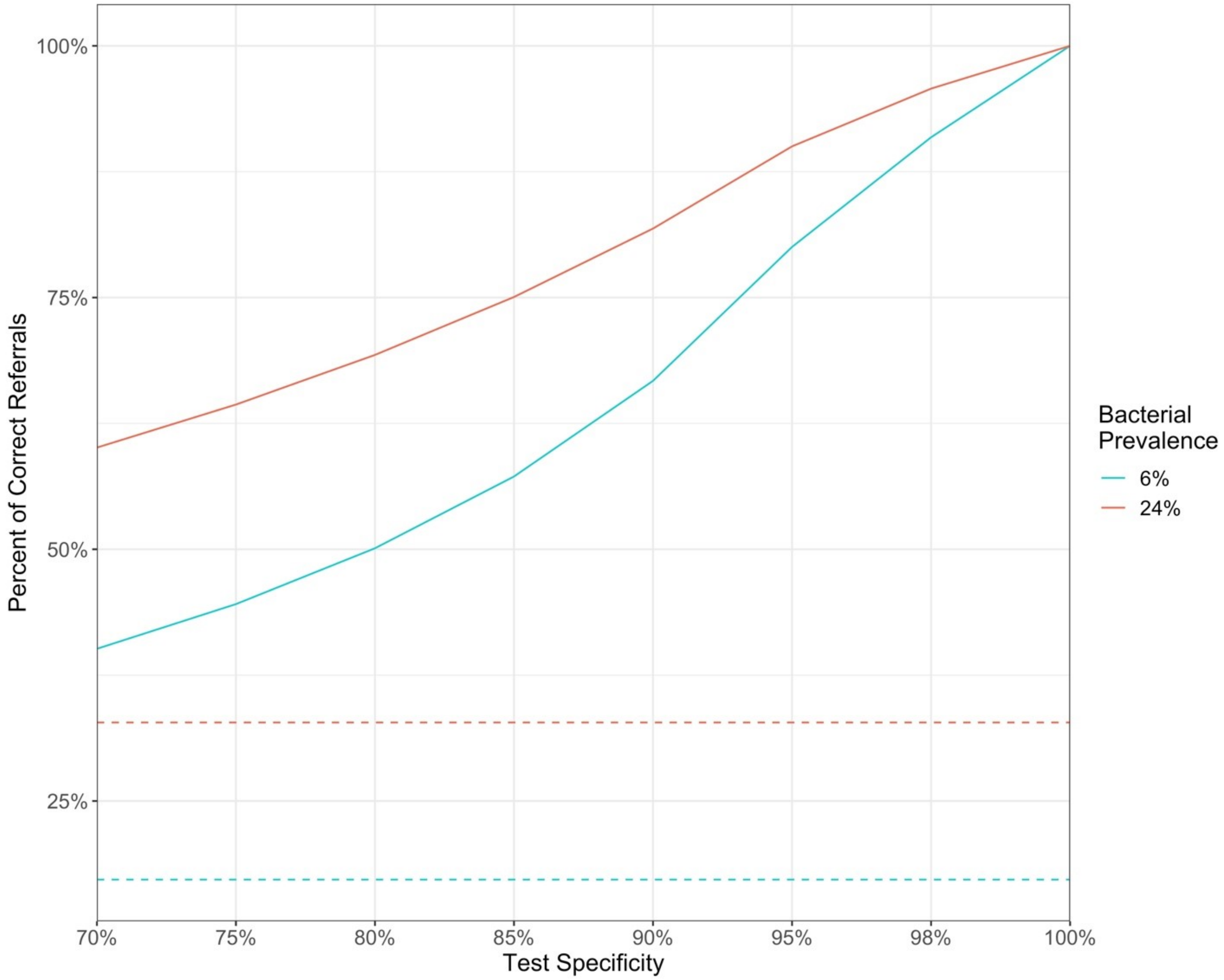

**Figure S5**. Total number of infant deaths at baseline per 1,000 possible serious bacterial infections in the community setting by bacterial prevalence, referral acceptance, hospital-associated infection risk, and hospital-associated infection case fatality rate. **HAI: hospital-associated infection, CFR: case fatality rate, pSBI: possible serious bacterial infection*.

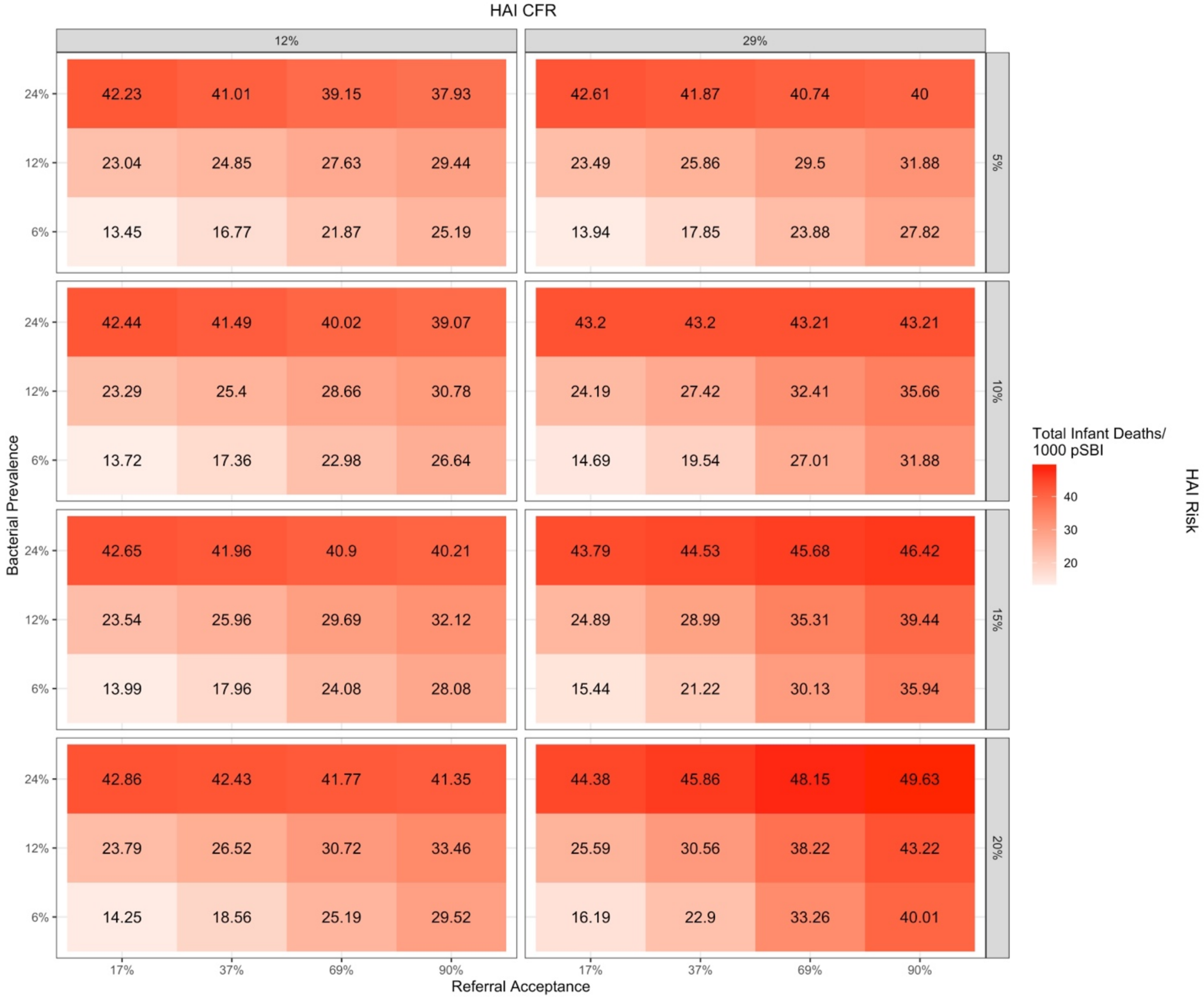

**Figure S6**. The percentage of community infant deaths at baseline attributable to hospital-associated infections, by bacterial prevalence, referral acceptance, hospital-associated infection risk, and hospital-associated infection case fatality rate. **HAI: hospital-associated infection, CFR: case fatality rate.*

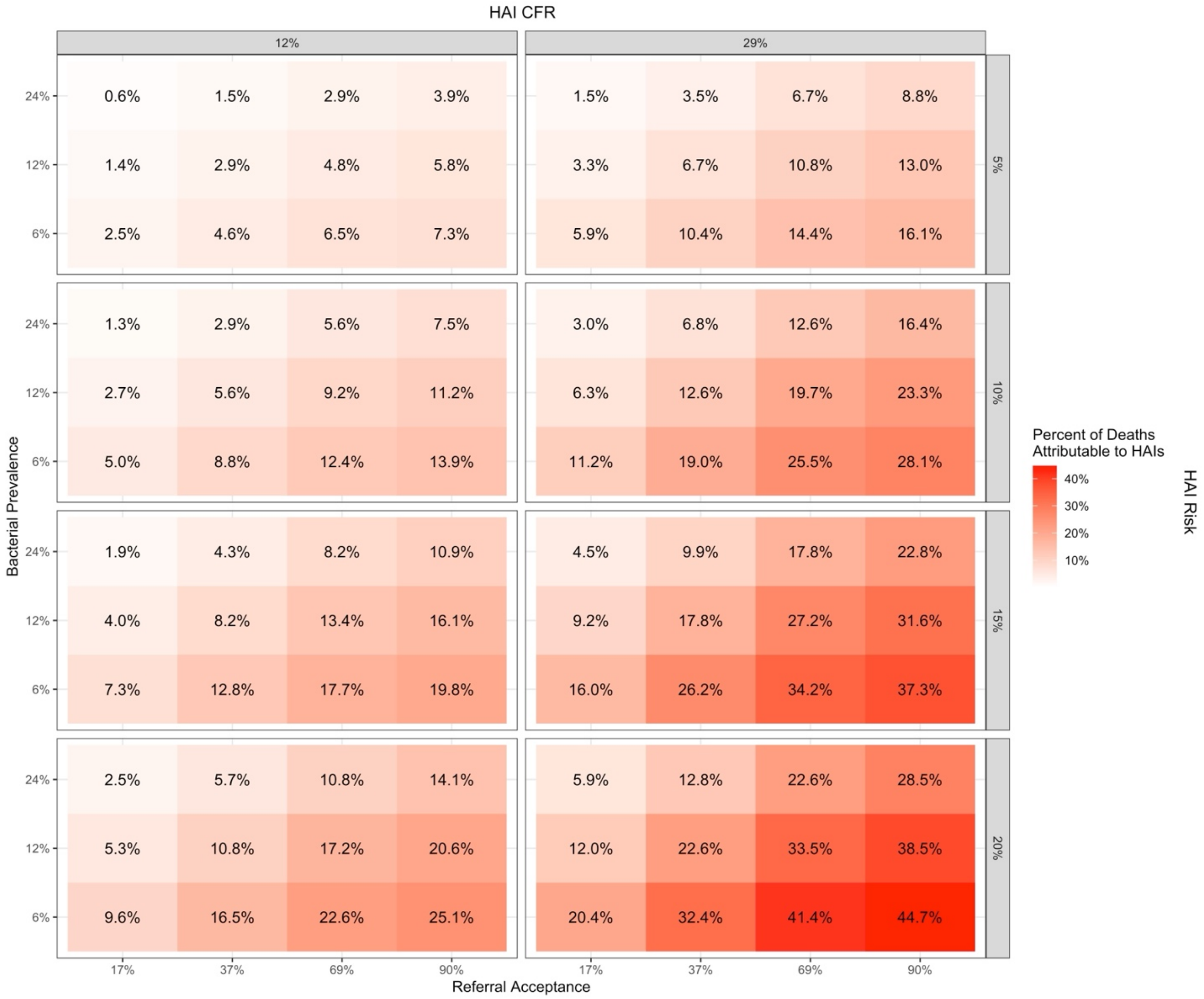

**Figure S7**. Percent reduction in hospital days compared to baseline when using the point-of-care test at the community level, by test sensitivity, specificity, bacterial prevalence, and referral acceptance. Hospital-associated infection risk is 20% and hospital-associated infection case fatality rate is 29%. **NICU: neonatal intensive care unit.*

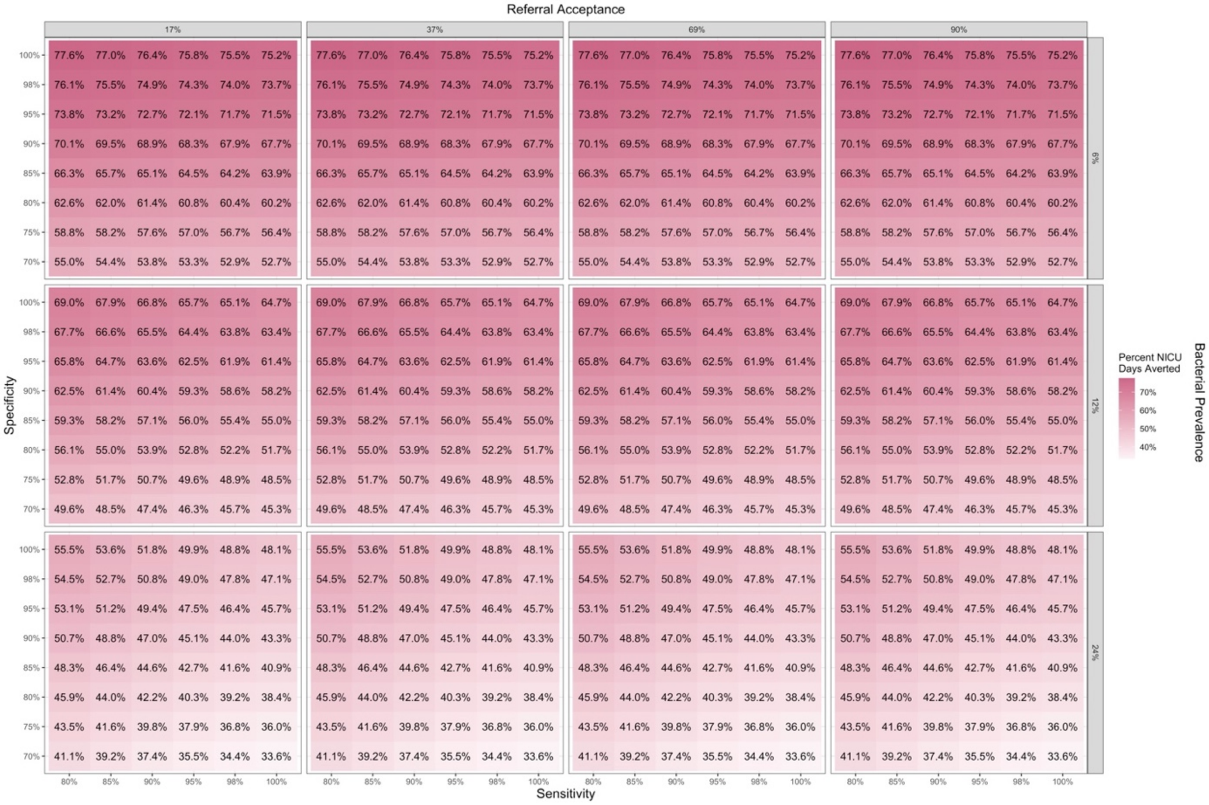

**Figure S8**. Threshold cost per point-of-care test in hospitals in India, by test sensitivity, specificity, bacterial prevalence, and referral acceptance. **A)** Hospital-associated infection risk is 5% and hospital-associated infection case fatality rate is 12%. **B)** Hospital-associated infection risk is 20% and hospital-associated infection case fatality rate is 12%. **HAI: hospital-associated infection*.

A)

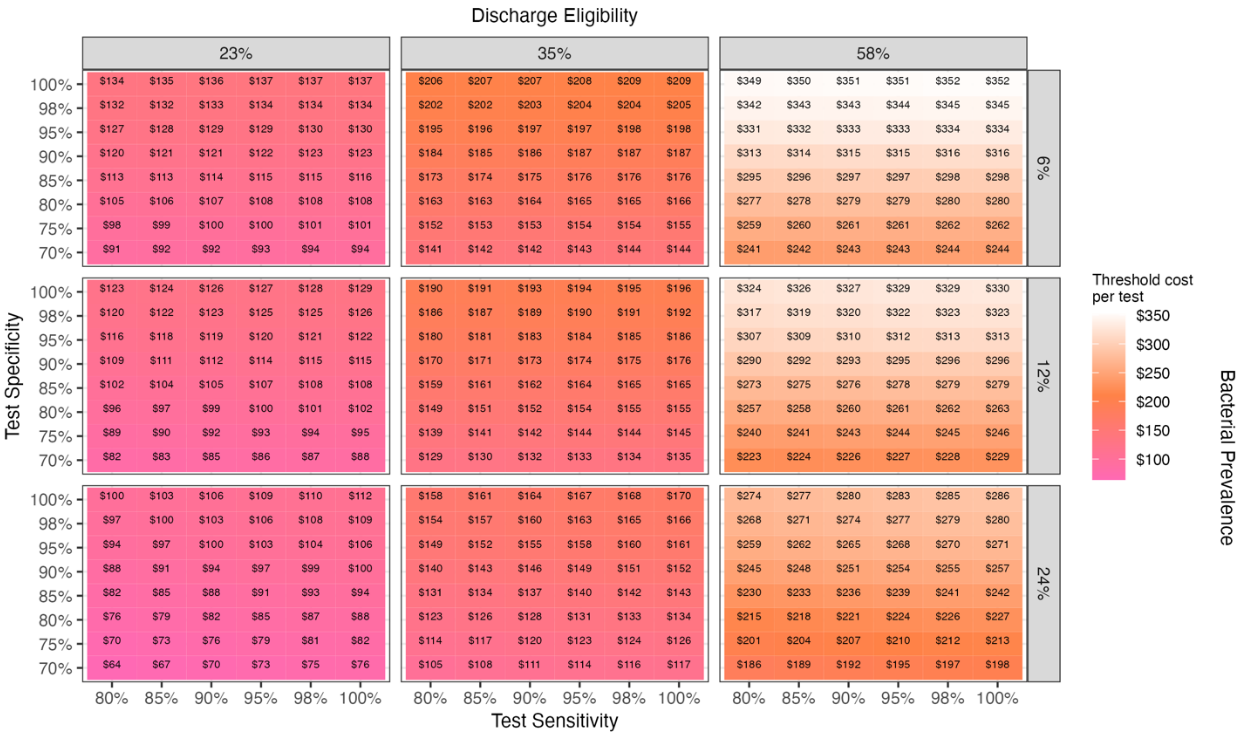

B)

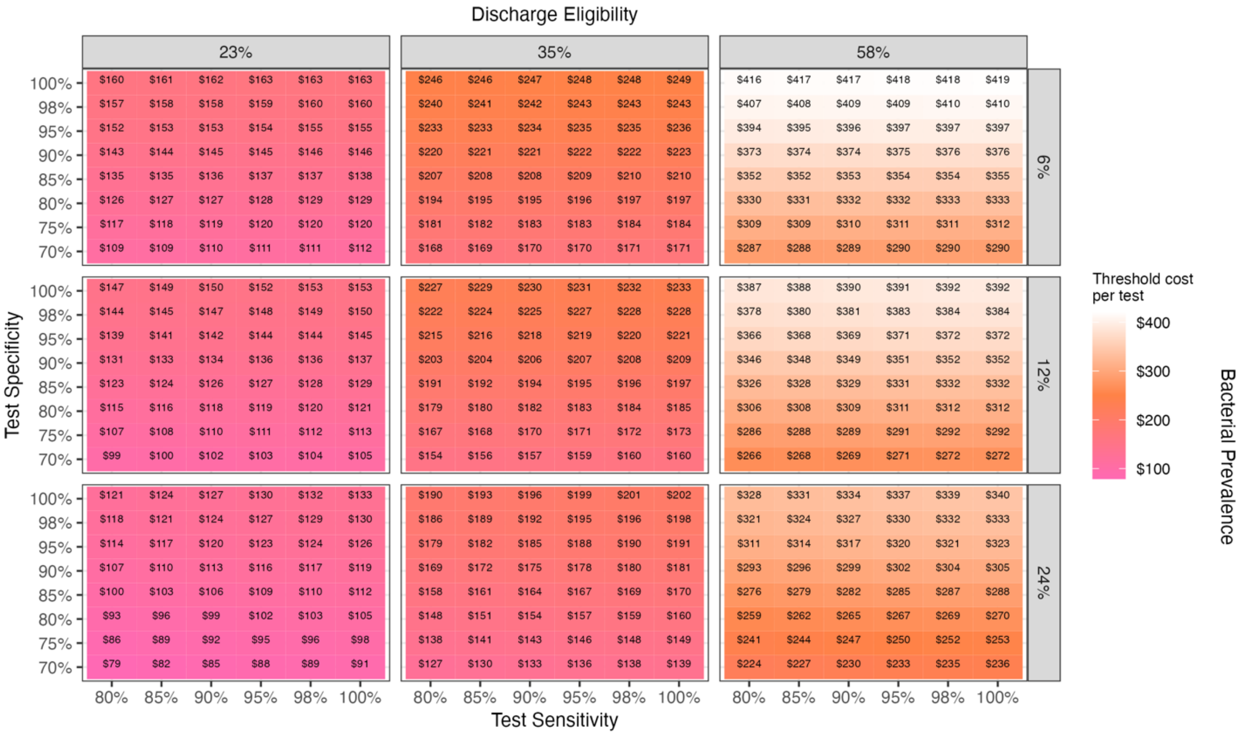

**Figure S9**. Threshold cost per point-of-care test in hospitals in Uganda, by test sensitivity, specificity, bacterial prevalence, and referral acceptance. **A)** hospital-associated infection risk is 5% and hospital-associated infection case fatality rate is 12%. **B)** hospital-associated infection risk is 20% and hospital-associated infection case fatality rate is 12%. **HAI: hospital-associated infection*.

A)

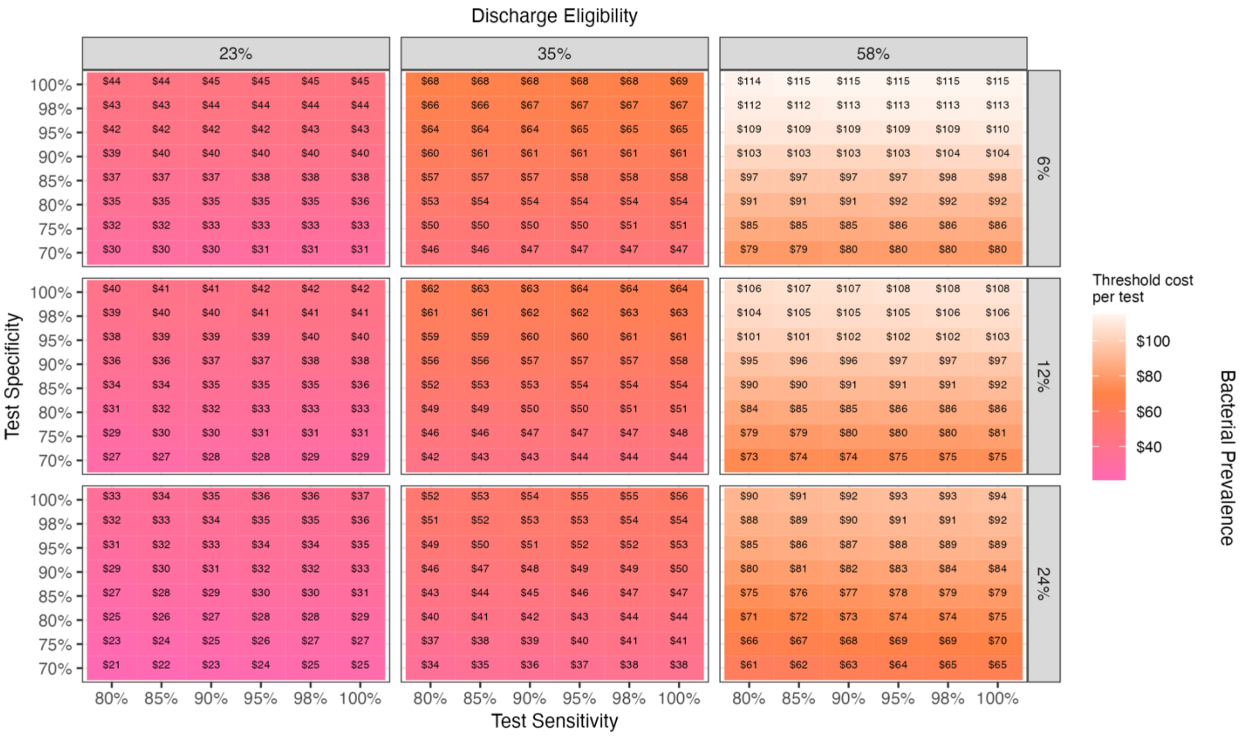

B)

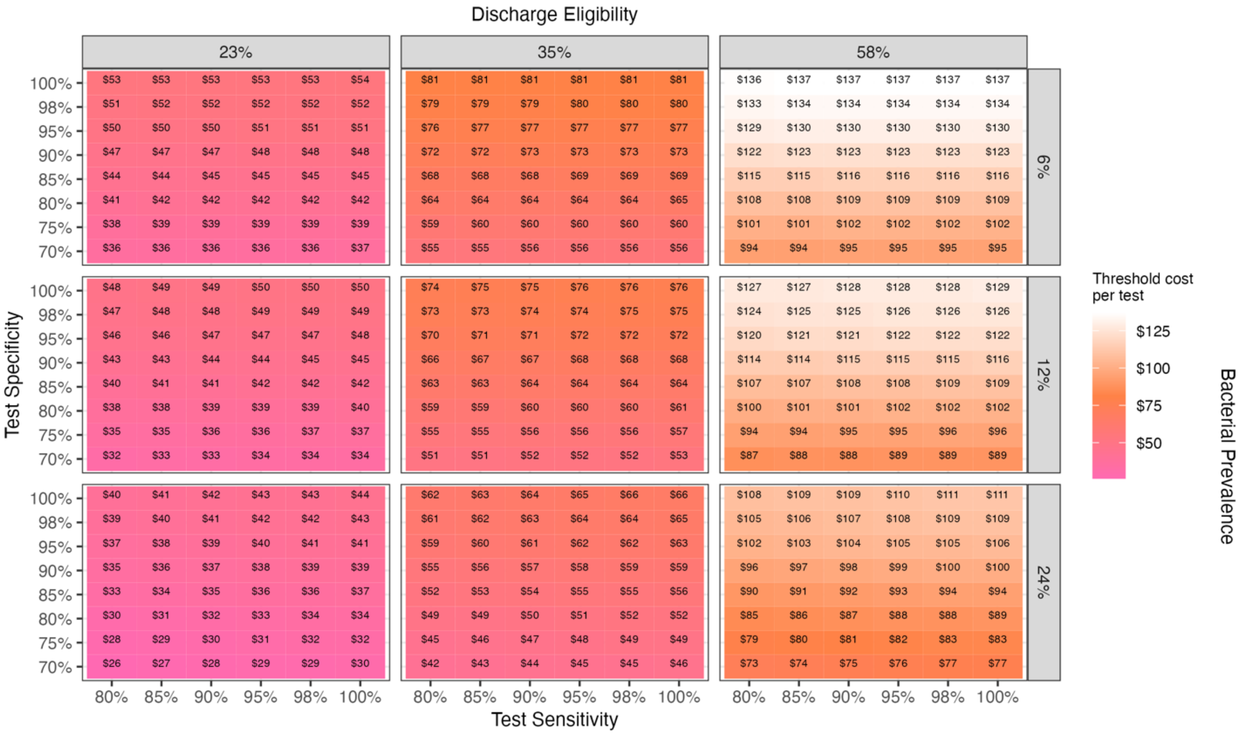

**Figure S10**. Threshold cost per point-of-care test at the community level in India, by test sensitivity, specificity, bacterial prevalence, and referral acceptance. **A)** Hospital-associated infection risk is 5% and hospital-associated infection case fatality rate is 12%. **B)** Hospital-associated infection risk is 20% and hospital-associated case fatality rate is 12%. **HAI: hospital-associated infection.*

A)

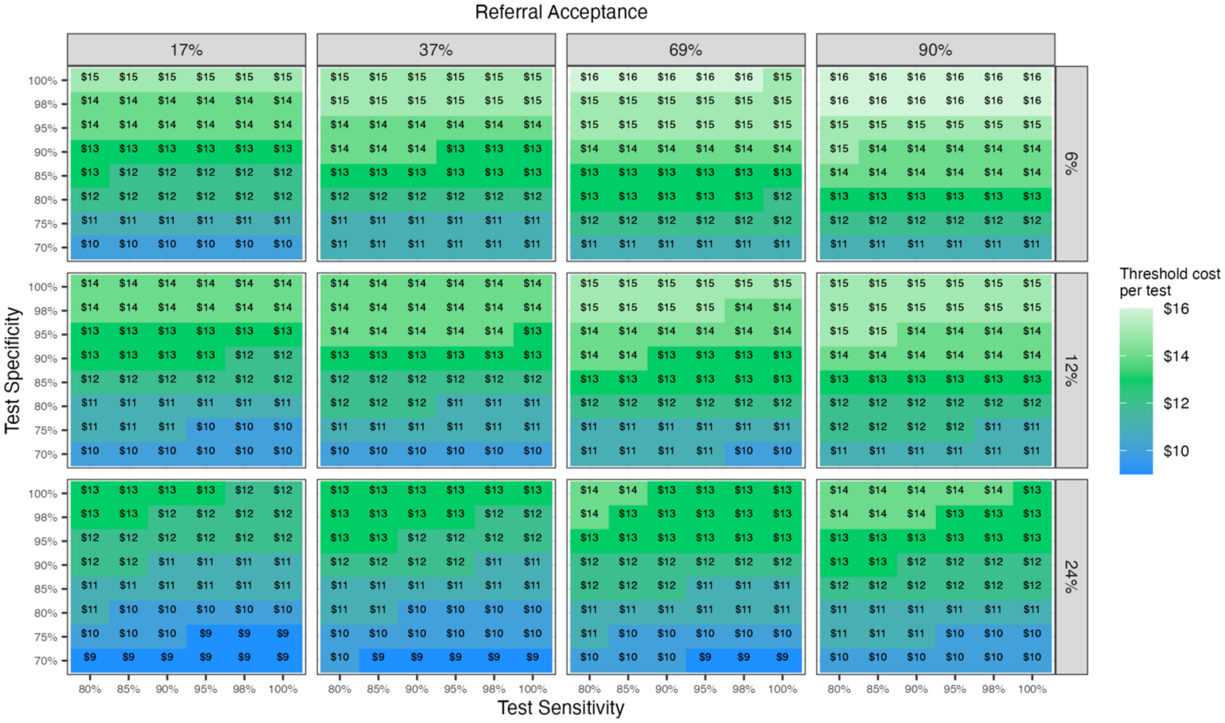

B)

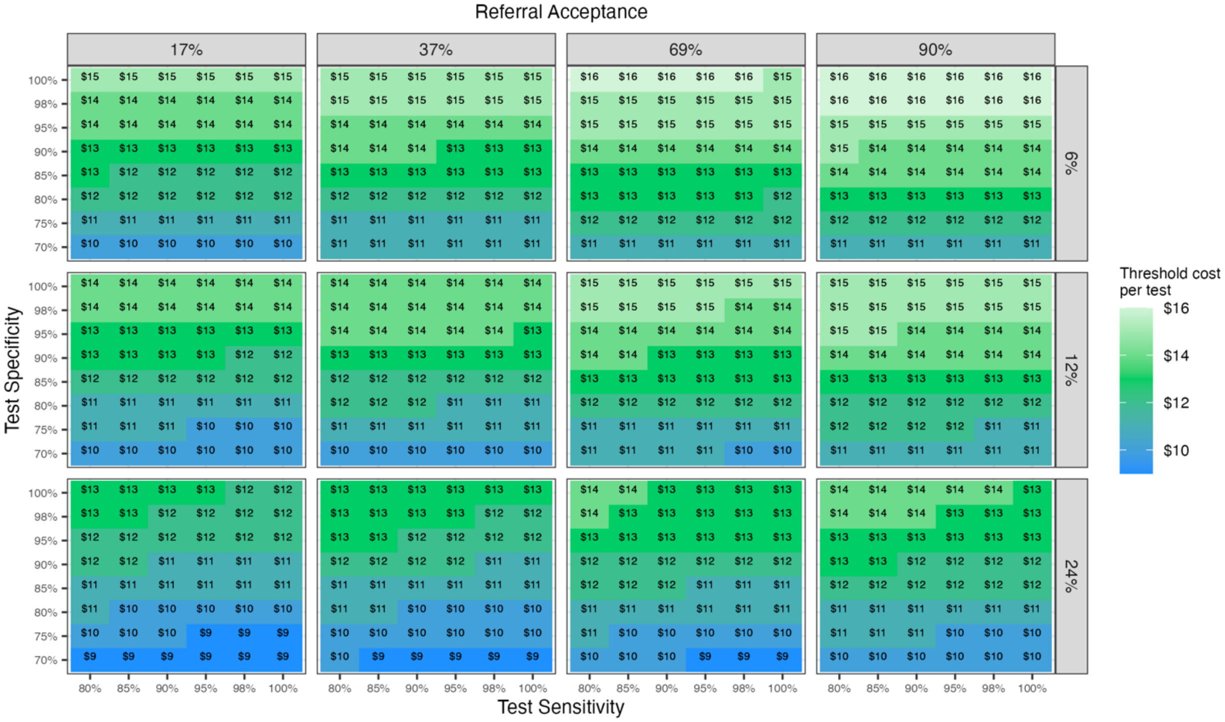

**Figure S11**. Threshold cost per point-of-care test at the community level in Uganda, by test sensitivity, specificity, bacterial prevalence, and referral acceptance. **A)** Hospital-associated infection risk is 5% and hospital-associated infection case fatality rate is 12%. **B)** Hospital-associated infection risk is 20% and hospital-associated case fatality rate is 12%. **HAI: hospital-associated infection*.

A)

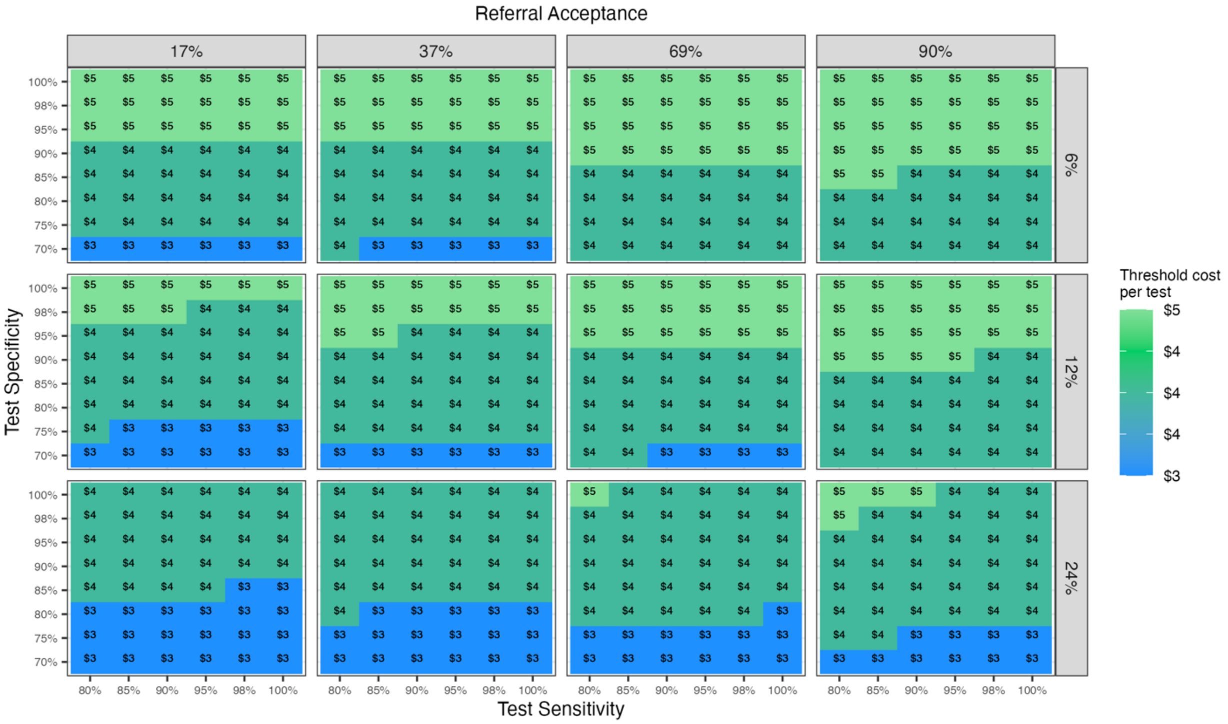

B)

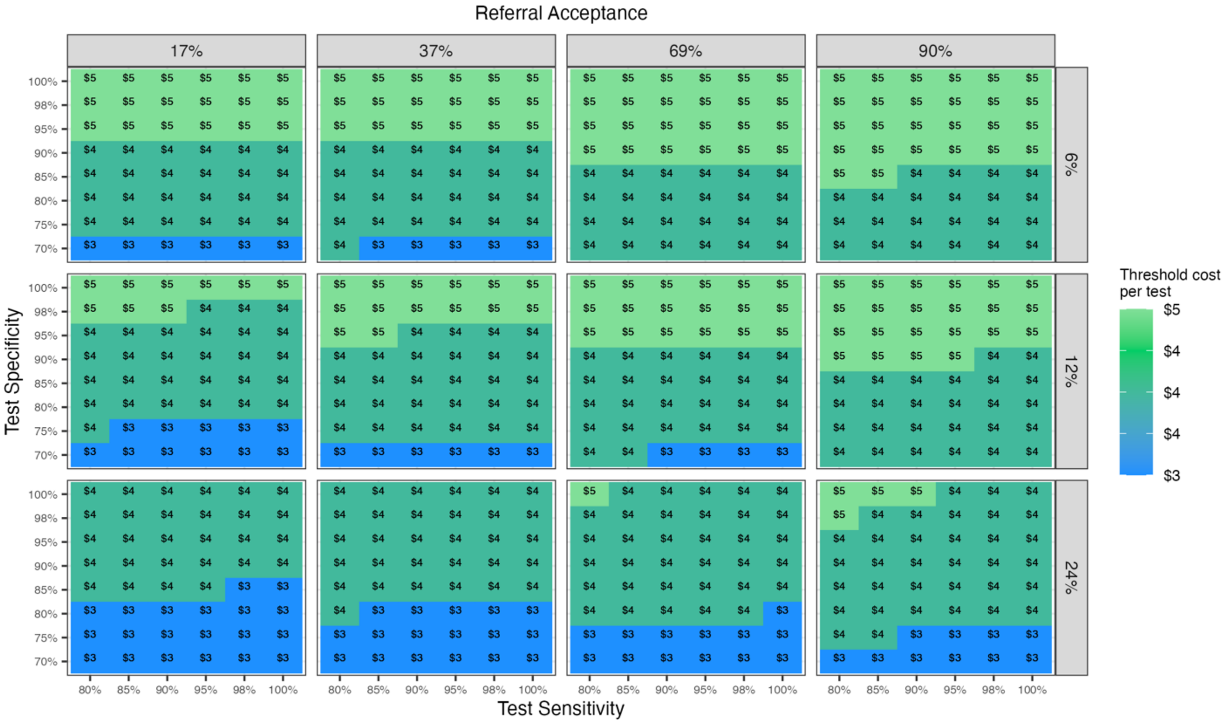

**Figure S12**. Threshold cost per point-of-care test at the community level (excluding infants with critical illness) in India, by test sensitivity, specificity, bacterial prevalence, and referral acceptance. **A)** Hospital-associated infection risk is 5% and hospital-associated infection case fatality rate is 12%. **B)** Hospital-associated infection risk is 20% and hospital-associated case fatality rate is 12%. **HAI: hospital-associated infection*.

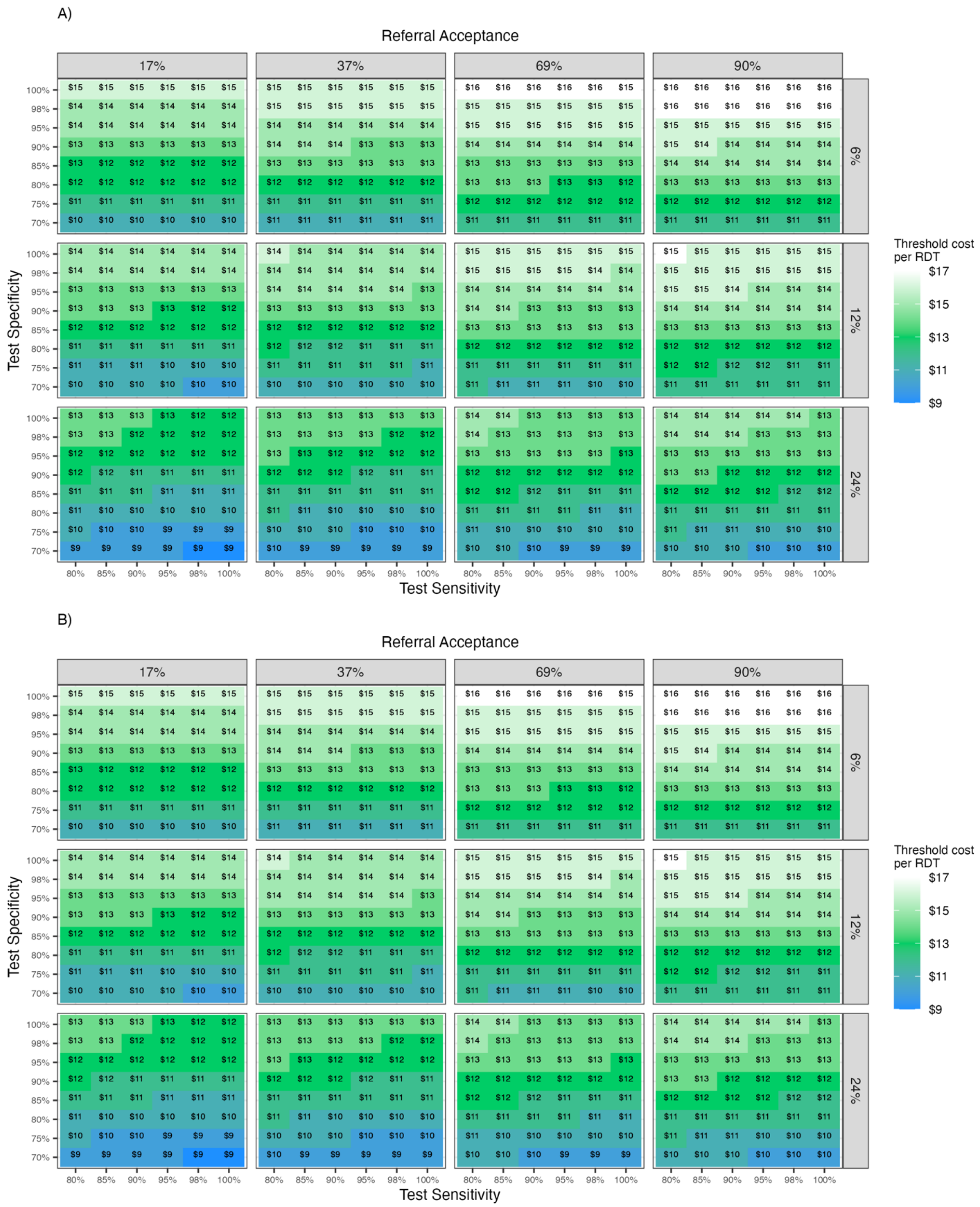

**Figure S13**. Threshold cost per point-of-care test at the community level (excluding infants with critical illness) in Uganda, by test sensitivity, specificity, bacterial prevalence, and referral acceptance. **A)** Hospital-associated infection risk is 5% and hospital-associated infection case fatality rate is 12%. **B)** Hospital-associated infection risk is 20% and hospital-associated case fatality rate is 12%. **HAI: hospital-associated infection*.

A)

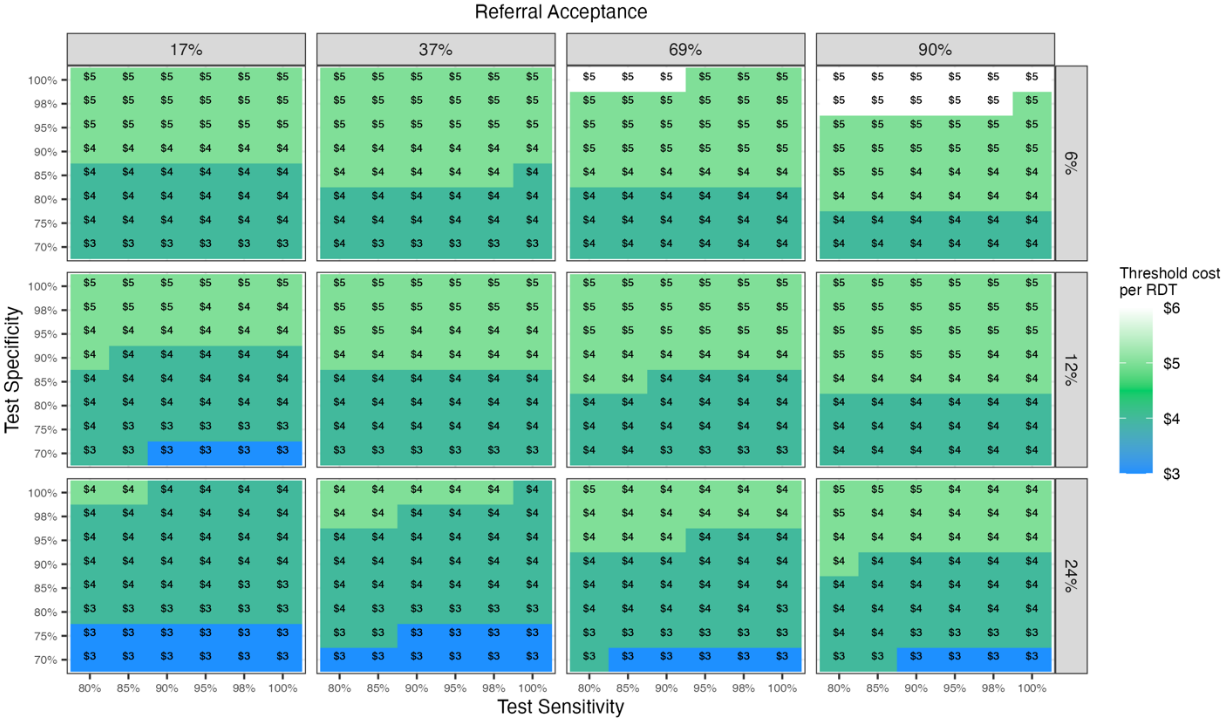

B)

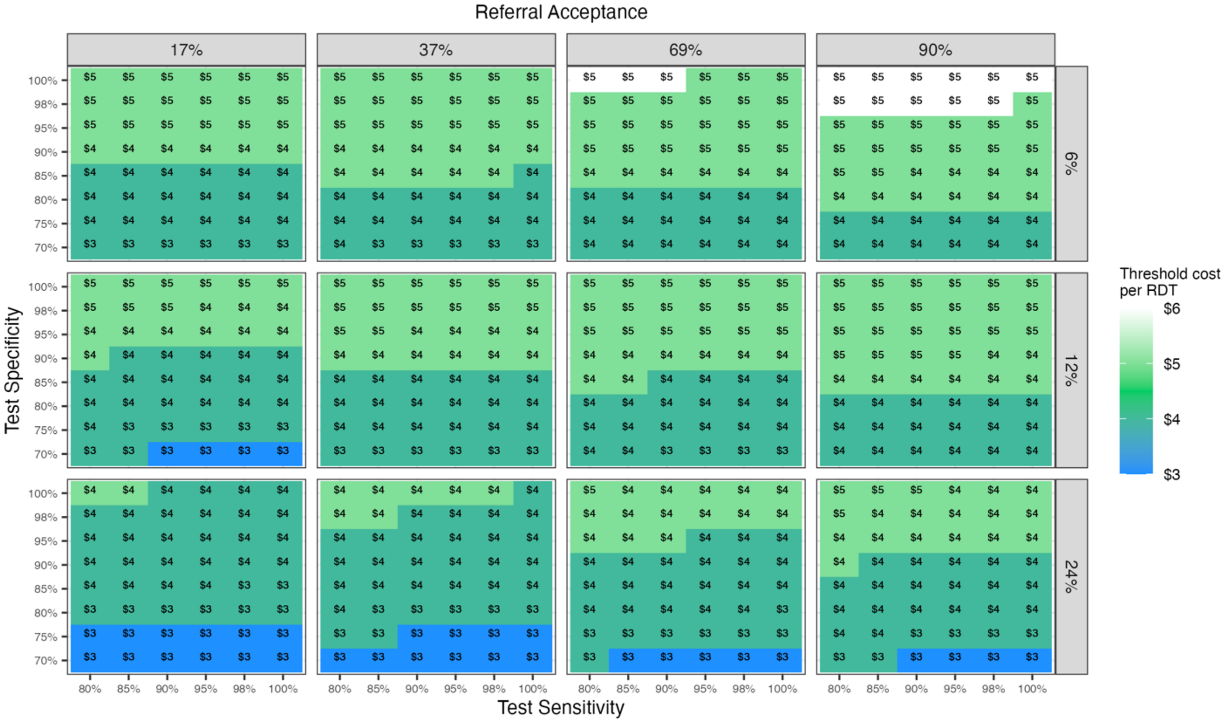

**Table S1.** Economic analysis determining the price of a point-of-care test (POCT) that maintains cost neutrality in India and Uganda in both a hospital and community setting, across a range of bacterial prevalence, percent of patients eligible for discharge, referral acceptance, hospital-associated-infection risk, and test sensitivity and specificity.

| Country | Bacterial Prevalence | Median POCT Cost | POCT cost range | |
|---|---|---|---|---|
| | | | Minimum | Maximum |
| **Hospitalized neonates** | | | | |
| **India** | **6%** | $196.94 | $90.84 | $418.74 |
| | **12%** | $183.47 | $81.97 | $392.49 |
| | **24%** | $155.54 | $64.24 | $339.98 |
| **Uganda** | **6%** | $64.56 | $29.78 | $137.28 |
| | **12%** | $60.15 | $26.87 | $128.67 |
| | **24%** | $50.99 | $21.06 | $111.46 |
| **Community-presenting infants** | | | | |
| **India** | **6%** | $13.43 | $10.22 | $16.03 |
| | **12%** | $12.81 | $9.71 | $15.38 |
| | **24%** | $11.58 | $8.70 | $14.08 |
| **Uganda** | **6%** | $4.42 | $3.36 | $5.27 |
| | **12%** | $4.21 | $3.19 | $5.06 |
| | **24%** | $3.81 | $2.86 | $4.63 |

**Table S2.** Economic analysis determining the price of a point-of-care test (POCT) that maintains cost neutrality in India and Uganda in the community setting (excluding infants with critical illness), across a range of bacterial prevalence, percent of patients eligible for discharge, referral acceptance, hospital-associated-infection risk, and test sensitivity and specificity.

| Country | Bacterial Prevalence | Median POCT Cost | POCT cost range | |
|---|---|---|---|---|
| | | | Minimum | Maximum |
| **Community-presenting infants *(excluding critically ill infants)*** | | | | |
| **India** | **6%** | $14.42 | $10.98 | $17.22 |
| | **12%** | $13.76 | $10.43 | $16.52 |
| | **24%** | $12.44 | $9.35 | $15.13 |
| **Uganda** | **6%** | $4.74 | $3.61 | $5.66 |
| | **12%** | $4.53 | $3.43 | $5.43 |
| | **24%** | $4.09 | $3.07 | $4.98 |

**Table S3.** CHEERS Checklist for Health Economic Evaluations[15]

| Topic | No. | Item | Location where item is reported |
|---|---|---|---|
| **Title** | 1 | Identify the study as an economic evaluation and specify the interventions being compared. | Title, page 1 |
| **Abstract** | 2 | Provide a structured summary that highlights context, key methods, results, and alternative analyses. | Abstract, page 2 |
| **Introduction** | | | |
| **Background and objectives** | 3 | Give the context for the study, the study question, and its practical relevance for decision making in policy or practice. | Introduction, page 4 |
| **Methods** | | | |
| **Health economic analysis plan** | 4 | Indicate whether a health economic analysis plan was developed and where available. | Methods, page 7 |
| **Study population** | 5 | Describe characteristics of the study population (such as age range, demographics, socioeconomic, or clinical characteristics). | Methods, pages 5-6 and Table 1 |
| **Setting and location** | 6 | Provide relevant contextual information that may influence findings. | Methods, pages 5-6 |
| **Comparators** | 7 | Describe the interventions or strategies being compared and why chosen. | Methods, pages 5-6 |

| | | | |
|---|---|---|---|
| **Perspective** | 8 | State the perspective(s) adopted by the study and why chosen. | Methods, page 7 |
| **Time horizon** | 9 | State the time horizon for the study and why appropriate. | Not applicable |
| **Discount rate** | 10 | Report the discount rate(s) and reason chosen. | Not applicable |
| **Selection of outcomes** | 11 | Describe what outcomes were used as the measure(s) of benefit(s) and harm(s). | Methods, page 7 |
| **Measurement of outcomes** | 12 | Describe how outcomes used to capture benefit(s) and harm(s) were measured. | Methods, page 7 |
| **Valuation of outcomes** | 13 | Describe the population and methods used to measure and value outcomes. | Methods, pages 5-7 and Table 1 |
| **Measurement and valuation of resources and costs** | 14 | Describe how costs were valued. | Methods, page 7 and Table 1 |
| **Currency, price date, and conversion** | 15 | Report the dates of the estimated resource quantities and unit costs, plus the currency and year of conversion. | Methods, page 7 and Table 1 |
| **Rationale and description of model** | 16 | If modelling is used, describe in detail and why used. Report if the model is publicly available and where it can be accessed. | Methods, pages 5-7 |

| | | | |
|---|---|---|---|
| **Analytics and assumptions** | 17 | Describe any methods for analyzing or statistically transforming data, any extrapolation methods, and approaches for validating any model used. | Methods, pages 5-7 |
| **Characterizing heterogeneity** | 18 | Describe any methods used for estimating how the results of the study vary for subgroups. | Methods, page 7 and Table 1 |
| **Characterizing distributional effects** | 19 | Describe how impacts are distributed across different individuals or adjustments made to reflect priority populations. | Methods, page 7 and Table 1 |
| **Characterizing uncertainty** | 20 | Describe methods to characterize any sources of uncertainty in the analysis. | Methods, page 7 and Table 1 |
| **Approach to engagement with patients and others affected by the study** | 21 | Describe any approaches to engage patients or service recipients, the general public, communities, or stakeholders (such as clinicians or payers) in the design of the study. | Not applicable |
| **Results** | | | |
| **Study parameters** | 22 | Report all analytic inputs (such as values, ranges, references) including uncertainty or distributional assumptions. | Methods, Table 1 |
| **Summary of main results** | 23 | Report the mean values for the main categories of costs and outcomes of interest and summarize them in the most appropriate overall measure. | Results, pages 10-12 |
| **Effect of uncertainty** | 24 | Describe how uncertainty about analytic judgments, inputs, or projections affect findings. Report the effect of choice of discount rate and time horizon, if applicable. | Methods, pages 10-12 |
| **Effect of engagement** | | | |

| | | | |
|---|---|---|---|
| **with patients and others affected by the study** | 25 | Report on any difference patient/service recipient, general public, community, or stakeholder involvement made to the approach or findings of the study | Not applicable |
| **Discussion** | | | |
| **Study findings, limitations, generalizability, and current knowledge** | 26 | Report key findings, limitations, ethical or equity considerations not captured, and how these could affect patients, policy, or practice. | Discussion, pages 13-15 |
| **Other relevant information** | | | |
| **Source of funding** | 27 | Describe how the study was funded and any role of the funder in the identification, design, conduct, and reporting of the analysis | page 16 |
| **Conflicts of interest** | 28 | Report authors conflicts of interest according to journal or International Committee of Medical Journal Editors requirements. | page 16 |
